# The association between mechanical ventilator availability and mortality risk in intensive care patients with COVID-19: A national retrospective cohort study

**DOI:** 10.1101/2021.01.11.21249461

**Authors:** Harrison Wilde, Thomas Mellan, Iwona Hawryluk, John M. Dennis, Spiros Denaxas, Christina Pagel, Andrew Duncan, Samir Bhatt, Seth Flaxman, Bilal A Mateen, Sebastian J Vollmer

## Abstract

**Objectives:** To determine if there is an association between survival rates in intensive care units (ICU) and occupancy of the unit on the day of admission.

**Design:** National retrospective observational cohort study during the COVID-19 pandemic.

**Setting:** 90 English hospital trusts (i.e. groups of hospitals functioning as single operational units).

**Participants:** 6,686 adults admitted to an ICU in England between 2^nd^ April and 1^st^ December, 2020 (inclusive), with presumed or confirmed COVID-19, for whom data was submitted to the national surveillance programme and met study inclusion criteria.

**Interventions:** N/A

**Main Outcomes and Measures:** A Bayesian hierarchical approach was used to model the association between hospital trust level (mechanical ventilation compatible) bed occupancy, and in-hospital all-cause mortality. Results were adjusted for unit characteristics (pre-pandemic size), individual patient-level demographic characteristics (age, sex, ethnicity, time-to-ICU admission), and recorded chronic comorbidities (obesity, diabetes, respiratory disease, liver disease, heart disease, hypertension, immunosuppression, neurological disease, renal disease).

**Results:** 121,151 patient-days were observed, with a mortality rate of 20.8 per 1,000 patient days. Adjusting for patient-level factors, mortality was higher for admissions during periods of high occupancy (>85% occupancy versus the baseline of 45 to 85%) [OR 1.18 (95% posterior credible interval (PCI): 1.00 to 1.38)]. In contrast, mortality was decreased for admissions during periods of low occupancy (<45% relative to the baseline) [OR 0.79 (95% PCI: 0.69 to 0.90)].

**Conclusion and Relevance:** Increasing occupancy of beds compatible with mechanical ventilation, a proxy for operational strain, is associated with a higher mortality risk for individuals admitted to ICU. Public health interventions (such as expeditious vaccination programmes and non-pharmaceutical interventions) to control both incidence and prevalence of COVID-19, and therefore keep ICU occupancy low in the context of the pandemic, are necessary to mitigate the impact of this type of resource saturation.

**Summary Box**

What is already known on this topic
Pre-pandemic, higher occupancy of intensive care units was shown to be associated with increased mortality risk. However, there is limited data on the extent to which occupancy levels impacted patient outcomes during the COVID-19 pandemic, especially in light of the mobilisation of significant additional resources. A recent study from Belgium reported a 42% higher mortality during periods of ICU surge capacity deployment, although in the analysis surge capacity was evaluated only as a binary variable, and notably this contradicts earlier results from smaller studies in Australia and Wales, where no association between ICU occupancy and mortality was identified.

What this study adds
The results of this study suggest that survival rates for patients with COVID-19 in intensive care settings appears to deteriorate as the occupancy of (surge capacity) beds compatible with mechanical ventilation (a proxy for operational pressure), increases. Moreover, this risk doesn’t occur above a specific threshold, but rather appears linear; whereby going from 0% occupancy to 100% occupancy increases risk of mortality by 69% (after adjusting for relevant individual-level factors). Furthermore, risk of mortality based on occupancy on the date of recorded outcome is even higher; OR 2.98 (95% posterior credible interval: 2.33 – 3.83). As such, this national-level cohort study of England provides compelling evidence for a relationship between occupancy and critical care mortality, and highlights the needs for decisive action to control the incidence and prevalence of COVID-19.

## Introduction

From the first reports of a novel coronavirus (SARS-CoV-2) in late 2019, to date, global mortality associated with the resultant disease (COVID-19) has exceeded 1.7 million people.[1] The virulence of the pathogen has prompted persistent concern about the ability of health services around the world to effectively care for the vast numbers of people affected.[2] These concerns are most relevant in the context of scarce resources (e.g., mechanical ventilation) required by patients in need of high-acuity support, which is relatively common in patients with COVID-19.[3] Notably, even with the introduction of non-pharmacological interventions such as stay-at-home orders, almost a third of all hospitals in England reached 100% occupancy of their “surge” mechanical ventilation capacity (i.e., including the additional beds that were created through the re-allocation of resources) during the first wave of the pandemic.[4] England is now experiencing a second wave that is already worse than the first, with 40% more people in hospital, many hospitals overwhelmed and exhausted staff.[5] What remains unclear is whether and to what extent operating at these extremes of critical care occupancy impacted patient outcomes.

Pre-pandemic, higher occupancy levels in intensive care (which may reflect operational strain), was shown to be associated with higher mortality risk.[6] However, there is limited data on the extent to which occupancy levels impacted patient outcomes during the first wave of COVID-19.[7] A recent study from Belgium reported 42% higher mortality during periods of ICU surge capacity deployment, although in the analysis surge capacity was evaluated only as a binary variable.[8] This contradicts earlier results from smaller studies in Australia and Wales, where no association between ICU occupancy and mortality was identified.[9,10] A better understanding of how operating under such extreme circumstances affects outcomes is crucial for two reasons: firstly, to allow hospitals to adapt practice to improve outcomes and secondly, to provide policy makers with more accurate information about the potential consequences of allowing health services to be overwhelmed. As such, in this study, we sought to evaluate the extent to which mortality risk in intensive care units (ICUs) over the course of the first wave of the COVID-19 pandemic in England could be explained by differences in occupancy.

## Methods

### Data Sources

Data on all intensive care unit (ICU) admissions across England were extracted from the COVID-19 Hospitalisation in England Surveillance System (CHESS).[11] Information on occupancy rates were extracted from the daily situation reports (i.e., ‘SitRep’).^4^ Both datasets are mandatory regulatory submissions for all National Health Service (NHS) acute care providers in England, and further details about them can be found in the eMethods.

### Study population

All ICU admissions reported to CHESS between 2^nd^ April – 1^st^ December (see eMethods for details on date selection), with presumed/confirmed COVID-19 (100% tested positive during their admission; see eMethods for details on diagnosis), aged 18 – 99, non-pregnant, and with valid admission and occupancy data, were eligible for inclusion (eFigure 1).

### Patient and Public Involvement

No patients were involved in the design, interpretation of the results, or dissemination of this study. However, we plan to disseminate the results to patients and the public in collaboration with several media partners whom have already agreed to report on this results.

### Recorded clinical features

#### Patient-Level Data

Information extracted from CHESS about each patient comprised: administrative features (admitting trust, admission date), demographic characteristics (age, sex, ethnicity), recorded comorbidities (obesity, diabetes, asthma, other chronic respiratory disease, chronic heart disease, hypertension, immunosuppression due to disease or treatment, chronic neurological disease, chronic renal disease, chronic liver disease). Ethnicity was coded as white, Asian (Subcontinent and other), black, mixed, and other; comorbidities were recorded as binary indicator variables, with missing entries assumed to reflect the absence of a comorbidity. The appropriateness of this assumption in CHESS has been previously explored.[12]

#### Occupancy Data

Trusts are groups of geographically co-located hospitals that function as a single organisational unit within the UK’s national healthcare system. Information extracted from SitRep about each trust comprised: pre-pandemic (January – March 2020) number of beds compatible with mechanical ventilation, the proportion of beds compatible with mechanical ventilation occupied on each day of the study period, and each trust’s geographical region.[4] Linkage was carried out based on the trust that an individual was admitted to and the date of ICU admission in CHESS; patients in CHESS were matched via their admission date to the relevant occupancy information from the corresponding date in SitRep.

#### Outcome

The primary outcome was in-hospital all-cause mortality; patients were followed up until death, discharge, transfer, or the final follow-up date of 22^nd^ December. Discharge and transfer were both treated as suggesting that the patient survived.

### Statistical Analysis

Descriptive summaries were generated as median and interquartile ranges for continuous variables, and frequency and percentage incidence for categorical factors. Exploratory analyses included: the relationship of the COVID-19 epidemic curve to bed occupancy at a national level (eFigure 2); the distribution of missingness amongst patients and trusts (eFigure 3); variation in age and comorbidity burden over the first wave (eFigure 4); the impact of modelling continuous variables either linearly, through the use of threshold functions, or via (standard cubic) splines/smooths (eMethods).

A Bayesian hierarchical approach was used to model the association between the trust, group and patient-level factors and mortality risk. Specifically, a generalised additive mixed model was utilised, with intercept and slope coefficients for population and group level effects, and a Bernoulli likelihood with logit function to link to mortality outcome. Coefficients were inferred by Markov chain Monte Carlo sampling, using Stan (CmdStan V2.25.0), with a model specified using BRMS (V2.14.4) in R (V4.0.3).[13-15] Further information on the Bayesian prior specification and modelling methods are reported in the eMethods.

As secondary analyses, two potential interactions were assessed: 1) baseline trust size and occupancy; 2) patient age and occupancy (results not reported due to insignificance). We also assessed the association of occupancy on the recorded outcome date with mortality, and occupancy expressed in terms of pre-pandemic ICU size. Several sensitivity analyses were carried out: 1) filtering for different degrees of missingness of patient-level comorbidity data at trust-level; 2) including all individuals still on the unit as of 22^nd^ December (and assuming they survive; n = 495); 3) including all individuals with a known outcome but no date whom were excluded in the data cleaning process (n = 8); 4) a random effect for all patient post 16^th^ June 2020 (i.e. the date of the Recovery Trials press-release regarding the effectiveness of dexamethasone); 5) adjusting for week of admission; 6) adjusting for trust and region as random effects; 7) additional patient-level factors: time from hospital admission to ICU admission, chronic liver disease and obesity; see eMethods for justifications.

## Results

6,686 individuals were included in this study following application of the inclusion/exclusion criteria (see eFigure 1), of whom 2,517 (37.6%) died. In total, 121,151 (median 12 days per patient; IQR: 6 – 23) patient-days were observed, equating to a mortality rate of 20.8 per 1,000 patient days. A full summary of the recorded patient-level characteristics is reported in Table 1.

**Table 1:**
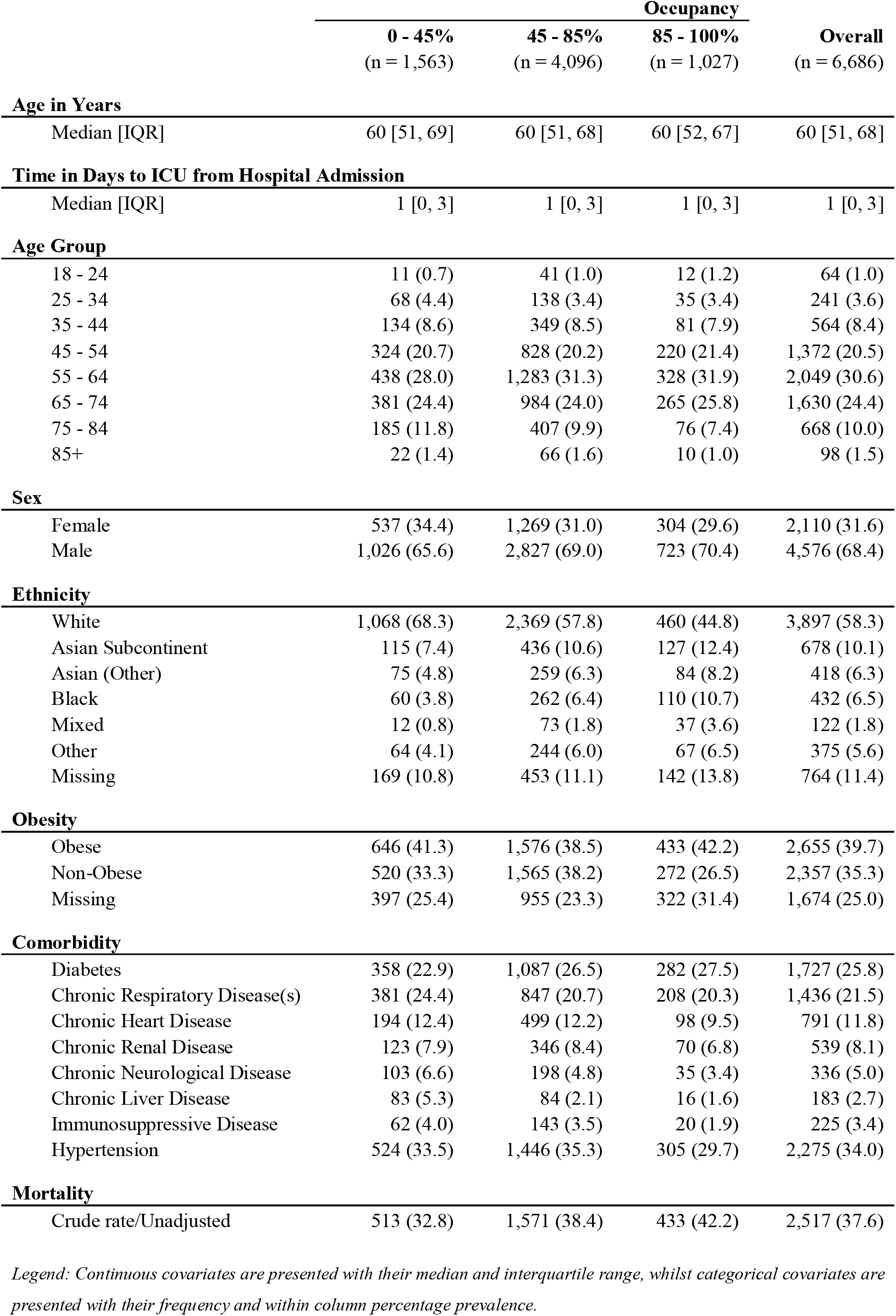
Characteristics of the study cohort stratified by occupancy on the day of admission.

|  | **Occupancy** |  |  | **Overall** |
| --- | --- | --- | --- | --- |
|  | **0 - 45%**<br>(n = 1,563) | **45 - 85%**<br>(n = 4,096) | **85 - 100%**<br>(n = 1,027) | **(n = 6,686)** |
| **Age in Years** |  |  |  |  |
| Median [IQR] | 60 [51, 69] | 60 [51, 68] | 60 [52, 67] | 60 [51, 68] |
| **Time in Days to ICU from Hospital Admission** |  |  |  |  |
| Median [IQR] | 1 [0, 3] | 1 [0, 3] | 1 [0, 3] | 1 [0, 3] |
| **Age Group** |  |  |  |  |
| 18 - 24 | 11 (0.7) | 41 (1.0) | 12 (1.2) | 64 (1.0) |
| 25 - 34 | 68 (4.4) | 138 (3.4) | 35 (3.4) | 241 (3.6) |
| 35 - 44 | 134 (8.6) | 349 (8.5) | 81 (7.9) | 564 (8.4) |
| 45 - 54 | 324 (20.7) | 828 (20.2) | 220 (21.4) | 1,372 (20.5) |
| 55 - 64 | 438 (28.0) | 1,283 (31.3) | 328 (31.9) | 2,049 (30.6) |
| 65 - 74 | 381 (24.4) | 984 (24.0) | 265 (25.8) | 1,630 (24.4) |
| 75 - 84 | 185 (11.8) | 407 (9.9) | 76 (7.4) | 668 (10.0) |
| 85+ | 22 (1.4) | 66 (1.6) | 10 (1.0) | 98 (1.5) |
| **Sex** |  |  |  |  |
| Female | 537 (34.4) | 1,269 (31.0) | 304 (29.6) | 2,110 (31.6) |
| Male | 1,026 (65.6) | 2,827 (69.0) | 723 (70.4) | 4,576 (68.4) |
| **Ethnicity** |  |  |  |  |
| White | 1,068 (68.3) | 2,369 (57.8) | 460 (44.8) | 3,897 (58.3) |
| Asian Subcontinent | 115 (7.4) | 436 (10.6) | 127 (12.4) | 678 (10.1) |
| Asian (Other) | 75 (4.8) | 259 (6.3) | 84 (8.2) | 418 (6.3) |
| Black | 60 (3.8) | 262 (6.4) | 110 (10.7) | 432 (6.5) |
| Mixed | 12 (0.8) | 73 (1.8) | 37 (3.6) | 122 (1.8) |
| Other | 64 (4.1) | 244 (6.0) | 67 (6.5) | 375 (5.6) |
| Missing | 169 (10.8) | 453 (11.1) | 142 (13.8) | 764 (11.4) |
| **Obesity** |  |  |  |  |
| Obese | 646 (41.3) | 1,576 (38.5) | 433 (42.2) | 2,655 (39.7) |
| Non-Obese | 520 (33.3) | 1,565 (38.2) | 272 (26.5) | 2,357 (35.3) |
| Missing | 397 (25.4) | 955 (23.3) | 322 (31.4) | 1,674 (25.0) |
| **Comorbidity** |  |  |  |  |
| Diabetes | 358 (22.9) | 1,087 (26.5) | 282 (27.5) | 1,727 (25.8) |
| Chronic Respiratory Disease(s) | 381 (24.4) | 847 (20.7) | 208 (20.3) | 1,436 (21.5) |
| Chronic Heart Disease | 194 (12.4) | 499 (12.2) | 98 (9.5) | 791 (11.8) |
| Chronic Renal Disease | 123 (7.9) | 346 (8.4) | 70 (6.8) | 539 (8.1) |
| Chronic Neurological Disease | 103 (6.6) | 198 (4.8) | 35 (3.4) | 336 (5.0) |
| Chronic Liver Disease | 83 (5.3) | 84 (2.1) | 16 (1.6) | 183 (2.7) |
| Immunosuppressive Disease | 62 (4.0) | 143 (3.5) | 20 (1.9) | 225 (3.4) |
| Hypertension | 524 (33.5) | 1,446 (35.3) | 305 (29.7) | 2,275 (34.0) |
| **Mortality** |  |  |  |  |
| Crude rate/Unadjusted | 513 (32.8) | 1,571 (38.4) | 433 (42.2) | 2,517 (37.6) |

*Legend: Continuous covariates are presented with their median and interquartile range, whilst categorical covariates are presented with their frequency and within column percentage prevalence.*

### Mechanical ventilator occupancy rate on the day of admission is associated with mortality risk

For high occupancy rates (85 – 100%), the unadjusted odds ratio (OR) for mortality based on the mechanical ventilator occupancy rate on the day of admission was 1.14 (95% posterior credibility interval (PCI): 1.00 – 1.33), relative to the baseline of 45 – 85%. For low occupancy rates (0 – 45%), the unadjusted odds ratio was 0.79 (95% PCI: 0.70 – 0.90), relative to the baseline of 45 – 85%. Following adjustment for patient-level factors (age, sex, ethnicity, and comorbidities), the OR was 1.18 (95% PCI: 1.00 – 1.38) for high occupancy rates, and 0.79 (95% PCI: 0.69 – 0.90) for low occupancy rates. Figure 1 illustrates the posterior probabilities for the fully adjusted ORs (see eFigure 5 & 6 for other model coefficients). To aid interpretation, the difference in risk for a 70-year-old man with no comorbidities being admitted during a period of high versus low occupancy is equivalent to the risk of them being over a decade older (Figure 2). Sensitivity analyses as detailed in eTable 1 were all concordant with the primary analysis.

**Figure 1:**
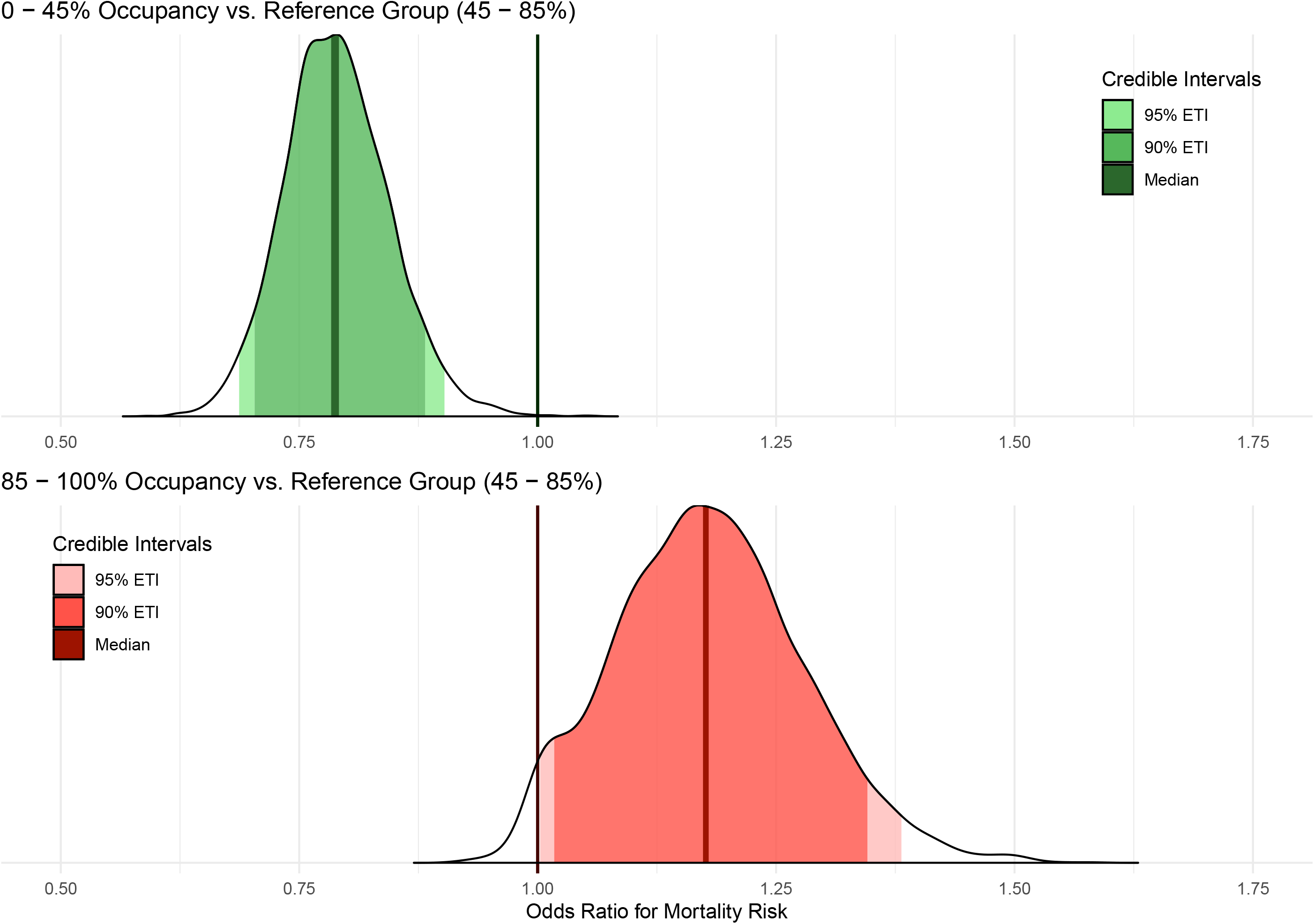
The adjusted odds ratios for the risk of mortality based on different ICU bed occupancy rates (treated as a three-level categorical variable) The full posterior distribution of the odds ratio (OR) for mortality given low occupancy 0 – 45% (Top; **Green**), and high occupancy 85 – 100% (Bottom; **Red**) are presented. The PCIs presented are equally tailed credibility intervals for the posterior odds ratio distributions. The outer (less saturated) interval is the 95% PCI, and the inner (more saturated) interval shows the 90% PCI. The reference category is 45 - 85% occupancy. Exact values are tabulated below.

**Figure 2:**
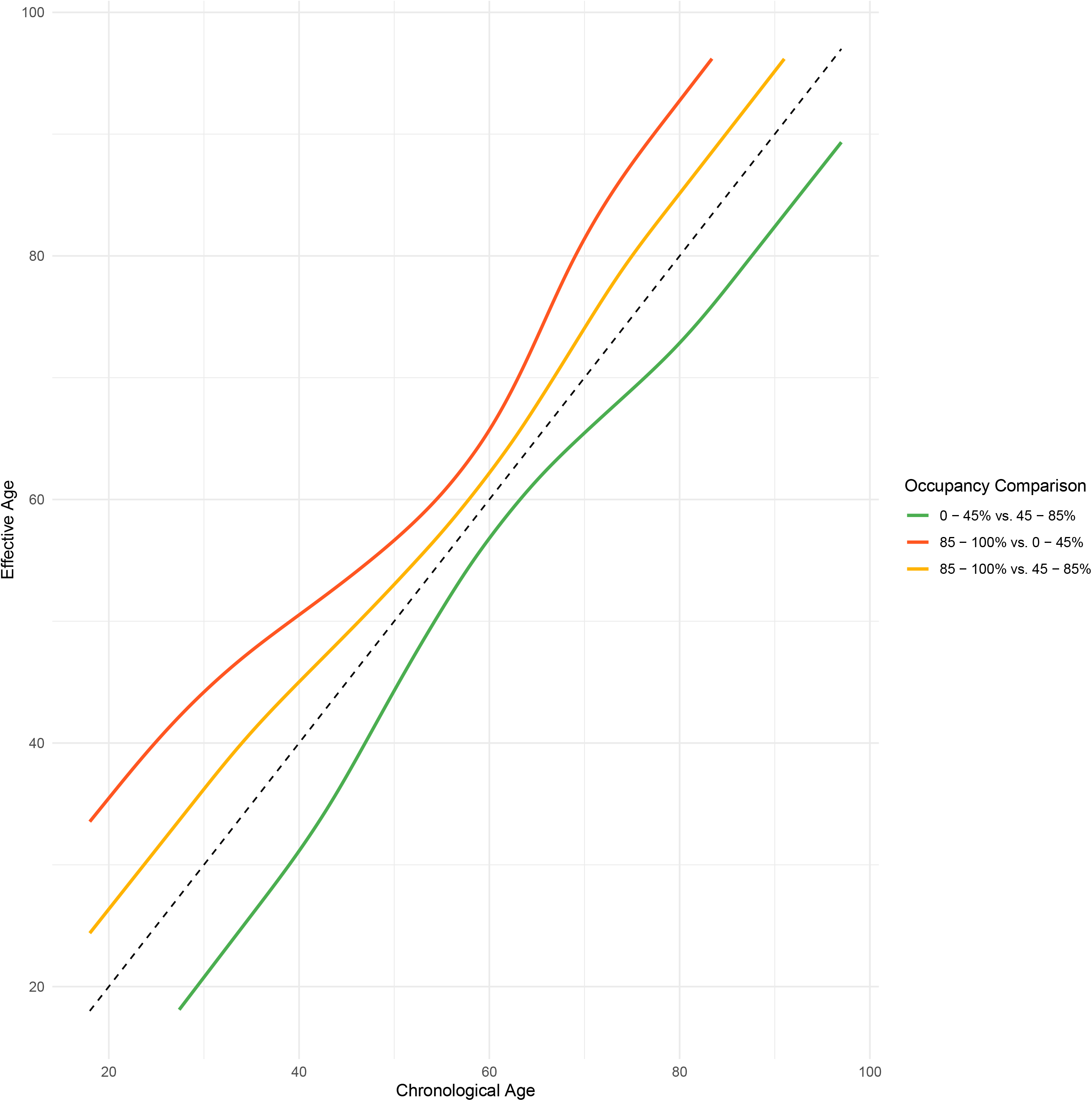
The increase in mortality risk associated with admission to intensive care during periods of different occupancy rates, expressed in terms of the equivalent increase in years of age. The plot illustrates the number of years of additional age that ICU admission on a day with each different mechanical ventilation bed occupancy rate equates to. For example, an individual with a chronological age of 40, has an effective age of 31 in a low occupancy setting (**Green**), and 45 in a high occupancy setting (**Orange**). Both of the aforementioned comparisons are relative to the baseline occupancy of 45 - 85%). A comparison of the difference in risk between being admitted to the highest occupancy range relative to the lowest is shown in (**Red)**, and for a 40-year-old is equivalent to an increase in their age by 11 years. The method for generating this specific type of plot is described in detail in the eMethods and via eFigure 7.[16]

### Mortality risk increases linearly with admission and date-of-outcome specific occupancy

The fully adjusted OR for mortality (Figure 3), using occupancy on the day of admission coded as a continuous linear variable ranging from 0 to 1 was 1.69 (95% PCI: 1.32 – 2.18). Moreover, using the bed occupancy from each individuals’ outcome date identified an even larger association (full model specification in eTable 2), OR 2.98 (95% PCI: 2.33 – 3.83).

**Figure 3:**
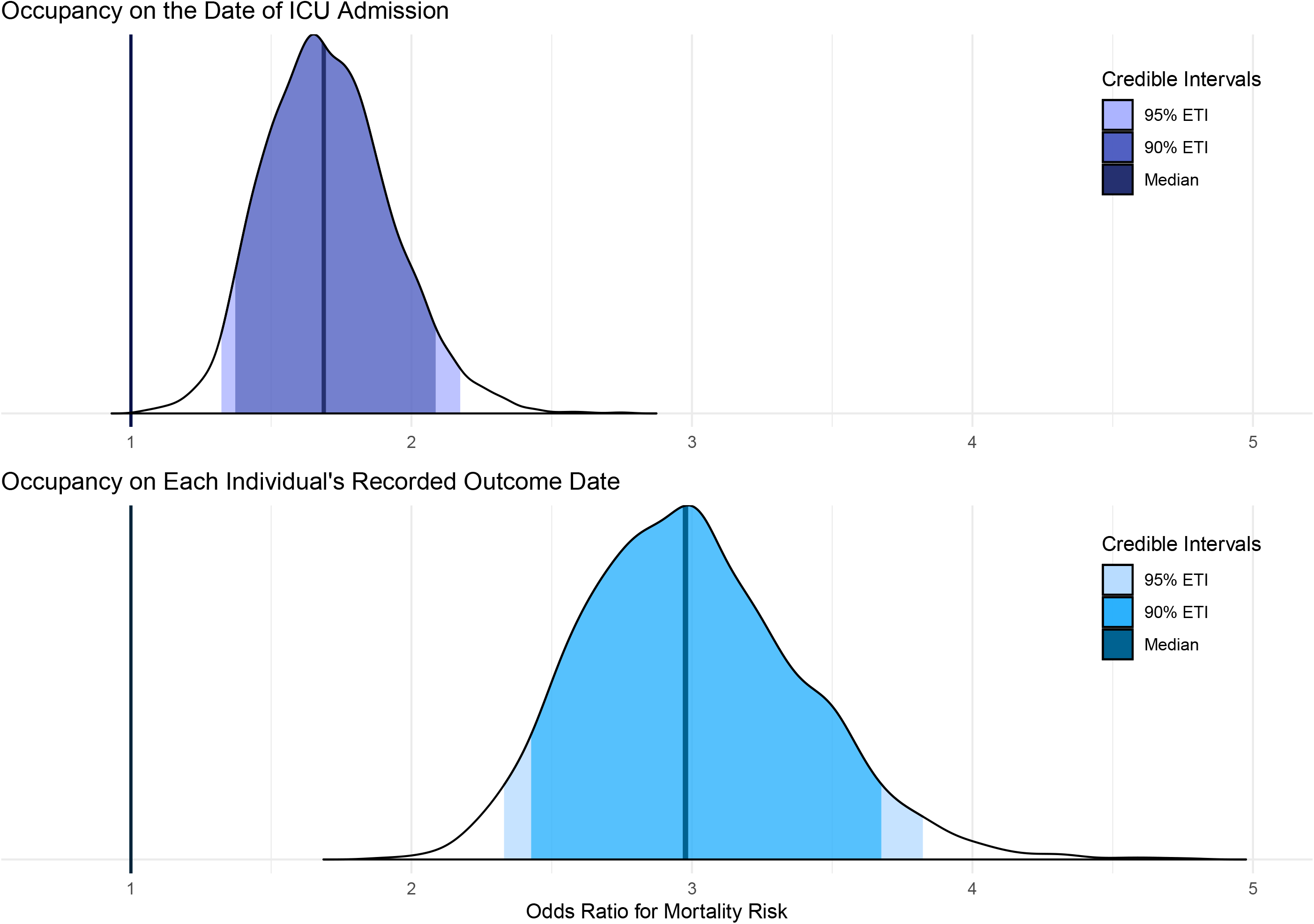
The adjusted odds ratios for the risk of mortality based on ICU bed occupancy (treated as a linear continuous variable) on the day of admission (top) and each individual’s recorded outcome date (bottom) The full posterior distribution of the odds ratio (OR) for mortality given occupancy on the date of ICU admission (Top; Purple), and occupancy on the date of each individual’s recorded outcome (Bottom; Blue) are presented. The PCIs presented are equally tailed credibility intervals for the posterior odds ratio distributions. Occupancy was specified without multiplying out by 100 (i.e., 20% = 0.20), therefore the odds ratio is for a step from 0% to 100% (i.e., 0.0 to 1.0). Exact values are tabulated below.

### Larger ICUs experience exaggerated impacts of extremes of mechanical ventilator occupancy rates

Although pre-pandemic number of beds did not substantially alter the OR of occupancy as a sensitivity analysis (eTable 1), it did appear independently informative. An increase in pre-pandemic ICU size by 25 beds compatible with mechanical ventilation was associated with a 22% decrease in risk of mortality (eTable 3). The introduction of an interaction term between pre-pandemic size and occupancy identified that larger ICUs experience exaggerated impacts of extremes of mechanical ventilator occupancy rates (eFigure 8).

## Discussion

The results of this study highlight a potential major impact of operational pressure on patient survival during the COVID-19 pandemic. Survival rates for patients with COVID-19 in intensive care settings appears to deteriorate as the occupancy of beds compatible with mechanical ventilation (a proxy for operational pressure),[17] increases. These observations are consistent with the aforementioned Belgian study,[8] except that our results suggest a linear association rather than single step increase at a specific threshold. Moreover, our findings also corroborate previous reports of an association between larger ICUs and lower COVID-19 mortality.[18] However, we additionally observe an interaction with (pre-pandemic) unit size, whereby larger ICUs experience more exaggerated impacts from both higher and lower (surge) occupancy rates. Although, It is unclear from our data what is driving this heterogeneity. Finally, these results might partially explain the decreased mortality rate seen in the latter half of the first wave in the UK,[19] where occupancy rates were much lower than at the peak.[4] Importantly, the average number of co-morbidities per patient appears to rise when occupancy decreases (eFigure 4), suggesting if there was implicit/soft triaging it would not have been in a manner that would explain the observed effect (i.e. more co-morbidities are generally thought to be associated with a worse outcome). However, more research is necessary to definitively exclude this potential explanation for the results.

### Strengths and Limitations

The strengths of this study are the national cohort of patient-level data with extensive capture of admissions,[20] coupled with a rigorous modelling method (eTable 4 & eMethods). Limitations include a lack of physiological data, limiting our ability to adjust for differences in severity upon admission. However, it is worth noting that previous studies using linked CHESS data from the first wave of the pandemic did not find between centre variation in severity scores (e.g. mean APACHE-II) to be associated with mortality risk.[21] Moreover, the characterisation of operational strain as a function of surge occupancy likely fails to fully reflect the complexity of using non-specialist staff and other resource allocation issues present when ‘creating’ new ICU beds (see eTable 5 using an alternative definition of occupancy based on baseline capacity; mortality risk given this linear continuous factor was 1.27 (95% PCI: 1.13 – 1.43). Finally, we lack clear 30-day outcome data for discharged and transferred individuals, and thus were forced to model under a naïve assumption that these individuals survived, which may have impacted our estimates of the risk.

### Implications for Policy Makers and Clinicians

In summary, our study highlights the importance of public health interventions (such as expeditious vaccination programmes and non-pharmacological interventions), to control both incidence and prevalence of COVID-19, and therefore actively manage ICU occupancy, as there is evidence of direct harm to patients as a consequence of saturation. This is especially relevant given the identification of a new strain of COVID-19 with a potentially increased risk of transmission,[22] coupled with observations that second wave-related operational pressures (including bed occupancy rates) in England have exceeded levels seen during the first wave of the pandemic,[5] suggesting immediate and decisive action is necessary.

## Supporting information

eMethods

## Data Availability

Data cannot be shared publicly as it was collected by Public Health England (PHE) as part of their statutory responsibilities, which allows them to process patient confidential data without explicit patient consent. Data utilised in this study were made available through an agreement between the University of Warwick and PHE. Individual requests for access to CHESS data are considered directly by PHE (contact via).

## Acknowledgements

HW is supported by the Feuer International Scholarship in Artificial Intelligence. TAM, IH and SB acknowledge funding from the MRC Centre for Global Infectious Disease Analysis (reference no. MR/R015600/1), jointly funded by the UK Medical Research Council (MRC) and the UK Foreign, Commonwealth & Development Office (FCDO), under the MRC/FCDO Concordat agreement and is also part of the EDCTP2 programme supported by the European Union. SB and SF were support by the MRC to undertake research on COVID-19 (MR/V038109/1). SB acknowledges funding from The Academy of Medical Sciences (SBF004/1080), Bill & Melinda Gates Foundation (CRR00280) and Imperial College Healthcare NHS Trust - BRC Funding (RDA02). JMD is supported by an Independent Fellowship funded by Research England’s Expanding Excellence in England (E3) fund. SJV, and BAM are supported by The Alan Turing Institute (EPSRC grant EP/N510129/). SJV is also supported by the University of Warwick IAA funding. SF is supported by EPSRC (EP/V002910/1). AD is supported by Wave 1 of the UKRI Strategic Priorities Fund under the EPSRC Grant EP/T001569/1.

## Contributors

BAM conceived the study, with input from SJV, and JMD. HW, JMD, and SJV prepared the data. HW, TM, and IH undertook the analysis, with input from SJV, SF, and BAM. All authors contributed to the development of the visualisations, and interpretation of results. BAM, HW, and TM drafted the manuscript. All authors critically revised the article and approved the final version for submission.

## Transparency Statement

The senior authors (SJV, BAM) had full access to all data and had final responsibility for the decision to submit for publication. SJV and BAM affirm that the manuscript is an honest, accurate, and transparent account of the study being reported; that no important aspects of the study have been omitted; and that any discrepancies from the study as originally planned have been explained.

## Ethics Approval

The study was reviewed and approved by the Warwick Biosciences Research Ethics Committee (BSREC 120/19-20-V1.1) and sponsorship is being provided by University of Warwick (SOC.28/19-20).

## Figure Legends

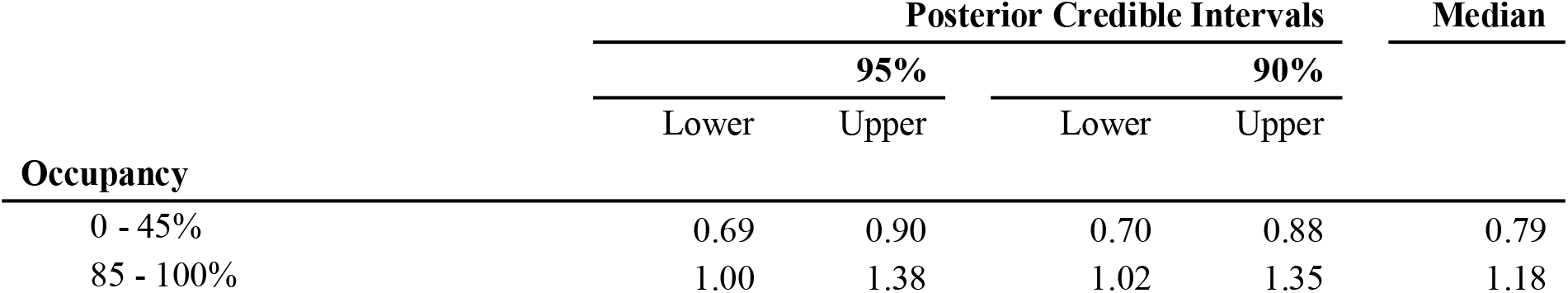

| Occupancy | Posterior Credible Intervals |  |  |  | Median |
| --- | --- | --- | --- | --- | --- |
|  | 95% |  | 90% |  |  |
|  | Lower | Upper | Lower | Upper |  |
| 0 - 45% | 0.69 | 0.90 | 0.70 | 0.88 | 0.79 |
| 85 - 100% | 1.00 | 1.38 | 1.02 | 1.35 | 1.18 |

| Occupancy | Posterior Credible Intervals |  |  |  | Median |
| --- | --- | --- | --- | --- | --- |
|  | 95% |  | 90% |  |  |
|  | Lower | Upper | Lower | Upper |  |
| Continuous Linear (Admission Date) | 1.32 | 2.18 | 1.37 | 2.09 | 1.69 |
| Continuous Linear (Outcome Date) | 2.33 | 3.83 | 2.42 | 3.68 | 2.98 |

