## Supplementary material for "The association between mechanical ventilator availability and mortality risk in intensive care patients with COVID-19: A national retrospective cohort study": eMethods

**eFigures (8)**

**eFigure 1**: Application of inclusion and exclusion criteria to raw data.

**eFigure 2**: Unadjusted mortality rate in the ICU, alongside the total, confirmed COVID and non-COVID national-level occupancies (Top) and percentages of the study cohort in each of the three main ICU occupancy bins used in the primary model (Bottom) across the duration of the study.

**eFigure 3**: Trust-level missingness at varying inclusion thresholds.

**eFigure 4**: Seven-day rolling averages for Age (Top) and Number of Comorbidities (Bottom) across the duration of the study.

**eFigure 5**: Marginal posterior densities for linear population level coefficients in the model, for mechanically ventilated bed occupancy binned as described in the main text, comorbidities, sex and week of admission.

**eFigure 6**: Marginal posterior densities for group level ethnicity intercepts.

**eFigure 7**: Predicted mortality risk for each of the three categorical levels of occupancy, illustrating how effective ages are calculated for Figure 2.

**eFigure 8**: The interaction between the baseline availability of beds supporting mechanical ventilation, and occupancy on the day of ICU admission.

**eTables (5)**

**eTable 1**: Marginal posterior densities for occupancy under sensitivity analyses.

**eTable 2**: Marginal posterior densities for mortality ORs when occupancy is utilised at each individual’s final outcome date, rather than their date of admission to ICU.

**eTable 3**: Marginal posterior densities for mortality ORs including baseline bed availability and interaction with occupancy.

**eTable 4**: Comparison of prior choices via their effect on primary model OR estimates.

**eTable 5**: Marginal posterior densities for mortality ORs when occupancy is defined as a ratio relative to baseline capacity.

**eMethods**

**Study Data**

*CHESS:* An earlier study by the UK’s intensive care audit group (ICNARC), suggested that a previous extract (from July 2020) accounted for 79% of ICU admissions over the associated period. The CHESS extract utilised in this study has nearly 900 more patients from the same period (added retrospectively due to delayed reporting), suggesting that as a result of this batch reporting, the most recent extract from CHESS has near-perfect capture of ICU admissions during the first wave of the pandemic (up to June 1^st^ 2020, when the British government formally started to wind down the first set of non-pharmaceutical interventions).[1] Moreover, comparisons to the corresponding demographic summaries and mortality statistics published by the audit team exhibits similar medians and distributions for the data present in both datasets, allowing for the fact that ICNARC includes data from February and March, which we do not.[2] For example, the mortality reported by ICNARC for this period was approximately 42.5%, whereas we found a slightly lower 37.6% (for the first wave of the pandemic), consistent with the fact that mortality rates slowly decreased over the first wave.[3] Similar data comparing the proportion of cases captured by CHESS for the summer and during the 2^nd^ set of non-pharmaceutical interventions introduced in mid-November until the 2^nd^ of December are not available, but in principle, we should expect less than perfect but still extensive capture (based on the experience of the 1^st^ wave). The CHESS extract utilised in this study was from the 4^th^ of January 2021. We stopped observing outcomes recorded after the 22^nd^ of December because this was when the first data on the new strain of COVID-19 was published by the British Government.[4] We hypothesized that this would fundamentally impact the natural experiment, and did not have data to control for this new variable. 40 people had outcomes recorded between the 22^nd^ of December and 4^th^ Jan (13 dead, 17 discharged, 10 transferred).

*SitRep*: This dataset includes total occupancy statistics per bed type, as well as COVID and Non-COVID specific occupancy breakdowns.[5] For this study we use the total occupancy statics. Importantly, the definition of a “utilised bed” by NHS-E and NHS-I for the SitRep is that of an “open and funded bed”. Effectively this requires adequate staff and resourcing for the bed to be considered operational, rather than just referring to a mattress or area in which a patient can be physically laid down. Analysis of CHESS data is limited to the intensive care settings, as mechanical ventilation is often only provided in these settings as per the UK’s definition of level 3 beds,[6] and as such this specific occupancy statistic in SitRep is ecologically relevant to the patient cohort we modelled. Furthermore, this flag is less prone to idiosyncrasies in organisation design where ICUs might include step-down HDU beds, which could manifest as double/inconsistent counting in SitRep. Finally, there is literature supporting the statement that the availability of these types of scare resources indicates operational pressure in ICUs.[7]

**Diagnostic Criteria**

As noted earlier, CHESS comprises both presumed and confirmed COVID-19 cases. The former were individuals with influenza-like infections (without a confirmatory diagnostic test) during the relevant study period. Data restricted to the first wave (i.e. up to June 1^st^ 2020, suggested at least 6% of people never tested positive, the more recent extract we have used suggested 100% of people had some form of positive test over the entire study period. During the first wave, diagnosis of COVID-19 was confirmed by reverse transcriptase polymerase chain reaction (PCR) of nasopharyngeal and/or oropharyngeal swabs or some of respiratory system washing/aspirate as it was the only available method during the study period.[8] However, national testing policy changed throughout the second half of study period, and it appears that CHESS began to include antigen and antibody tests. The testing principle was that all people who were admitted to ICU were tested, and people were potentially re-tested multiple times if the initial results were negative but clinical suspicion remained high. Moreover, previous sensitivity analysis has shown excluding groups that didn’t test positive based on a PCR test from the main analysis does not appear to impact association estimation, and thus we did not include this sensitivity analysis here.[9]

**Study Date Selection**

The time taken to set up the necessary data collection processes meant that CHESS data is only available from 1^st^ March 2020, and occupancy data for ICUs from 2nd April 2020. As such, the inclusion period was restricted to 2^nd^ April – 1^st^ December to ensure that occupancy on ICU admission date was available for all of the included patients. 2^nd^ December was the first working day following the English government’s announcement that the non-pharmacological interventions introduced in response to the pandemic would be de-escalated (at the end of the second lockdown), and thus is taken as the natural endpoint for the second wave. Moreover, the new, more transmissible, variant B 1.1.7 was formally acknowledged by the British government on the 22^nd^ of December,[4] and thus we halted all observations following this announcement as the natural experiment is likely to have been fundamentally impacted and we do not have the data to adjust for this confounder at present. We assess the impact of this incomplete episode observation via the aforementioned sensitivity analyses.

**Exploratory analysis**

As a post-hoc exploration, we also sought to describe variation in the case-mix to determine whether there was evidence of individuals with fewer comorbidities being admitted later in the first wave when occupancy was lower, which might have confounded the decreased mortality rate observed (eFigure 4).

**Primary Model Specification**

The primary model has population level coefficients for the following demographic features: *Age*, and *Sex*. Moreover, there are population level coefficients for the following comorbidities: *Diabetes*, *Chronic Respiratory Disease(s)*, *Immunosuppression*, *Chronic* *Renal Disease*, *Hypertension*, and *Chronic Heart Disease*. The final and primary feature of interest is regarding the proportion of beds supporting mechanical ventilation occupied, referred to simply as *Occupancy* and characterised in various ways discussed below.

*Age* is treated as a continuous variable and was re-parameterised using a cubic spline with 4 knots to investigate and subsequently represent non-linearity in the effect. *Sex* and the various comorbidity indicators are dichotomous. The *Chronic Respiratory Disease(s)* feature represents the union of two initially separate covariates in the data: *Respiratory Disease* and *Asthma*, setting instances where both are either missing or no to be negative; otherwise positive. *Ethnicity* was included in the model as a group level effect with an intercept coefficient for each of 7 groups: White, Asian Subcontinent, Asian (Other), Black, Mixed, Other and Missing.

For *Occupancy*, initially a binary variable indicating whether a patient entered an ICU with over 85% of beds occupied was formulated (this cut off is in accordance with guidelines from the Royal College of Emergency Medicine Guidelines).[10] This first attempt yielded a significant result (Median 1.26, 95% CI: 1.07 – 1.47). This was subsequently expanded into four “bins” with cut offs at 45%, 66.6% and 85% (the former two values split what remains of the data after the cut at 85% into 3 evenly sized groups based on quantiles), to explore whether there was further effect heterogeneity at lower occupancies. Here the 45 - 66.6% bin taken as a reference category left the 66.6 - 85% bin’s coefficient centred convincingly at 1 (Median 1.00, 95% CI: 0.89 – 1.11). Following this, the two middle bins were merged to result in the final formulation used in our primary analysis: a three-bin approach covering the ranges 0 – 45%, 45 – 85% and 85 – 100% bed occupancy. The occupancy ratio was also treated linearly and with a four-knot cubic spline; in both cases, the effect did appear significant and convincingly linear by visual inspection (Median 1.69, 95% CI: 1.32 – 2.18 for the linear OR estimates on occupancy ratio ranging from 0 to 1).

**Prior specification and checks**

To prevent overfitting, a hierarchical shrinkage (HS) prior was placed on population effects and smooths.[11] Whilst the HS prior encourages a sparse solution, we note that the predictors with the largest coefficients are the same as those arising from a standard Normal prior -- the main difference is that the HS prior enhances the concentration in posterior density at zero for coefficients that are small or weakly identified with Normal priors. The model coefficients inferred do not depend strongly on the effective degrees of freedom in the HS prior, which was tested for values between 1 and 7, nor was any degree of freedom parameter choice identifiably more appropriate based on Pareto-smoothed importance sampling leave-one-out (PSIS-LOO) cross validation.[12] Furthermore, PSIS-LOO cross validation was used to assess selection of the model structure - a more complex model with interactions between linear predictors was not justified. Results of prior predictive checks are given in eTable S5.

The primary model presented in this work has a sparsity promoting HS prior with 1 degree of freedom covering linear population effects and smooths to minimise overfitting, and a weakly informative student’s t (3, 0, 2.5) on the grouping of ethnicity intercepts. The prior predictive distributions generated by these choices span the range of plausible values. The marginal coefficient densities inferred were inspected for a range of prior choices - both with HS priors with differing degrees of freedom, and standard normal (N(0,1)) priors. It is reasonable to conclude that effect sizes from the model inference are insensitive to prior specification as shown in eTable 4; and whilst standard normal priors yield notably denser estimations compared to HS priors; the ordering of effect sizes is unchanged.

**Sampling Details**

The model parameter space was sampled using Hamiltonian Monte Carlo with 3 chains of 3,000 iterations each. The target proposal acceptance probability was modified from the default value to 0.95 to improve convergence with the hierarchical shrinkage prior. All models discussed had fewer than 1 in 1000 divergent transitions and R-hat diagnostics of 1.00. The minimum bulk effective sample size was 2000 for group coefficients and 3500 for population coefficients, and 3000 for smooth ones.

**Sensitivity Analyses**

Several sensitivity analyses were carried out, as noted in the main text. Specifically, filtering for different degrees of missingness at trust-level: by removing all trusts from the modelling data with a total of 25% of all comorbidity information missing, or 50%, or 75%. Moreover, adjustment for week of admission was carried out using a continuous term that was explored for evidence of non-linearity using a cubic spline with 4 knots. The results of the exploration were consistent with a linear feature (based on visual inspection and model comparison), and thus it was treated as such. Notably, this is consistent with prior analyses of similar data.[3] Adjustments for trust and region as random effects were undertaken. Furthermore, a set of additional patient-level factors (held out from the primary analysis) were adjusted for, including: time from hospital admission to ICU admission, chronic liver disease (CLD) and obesity. Previous studies from the UK suggested that the former (time to ICU admission) is unlikely to be informative, and so we planned for a more parsimonious primary model based on these results but wanted to ensure we identified a similar lack of effect. The latter two were not utilised in the primary model because the association between CLD and COVID-19 mortality is unclear,[13,14] and the degree of missingness in obesity makes it liable to mis-specify other associations.[15] Finally, sensitivity analyses were carried out surrounding the final outcomes associated with each patient. The primary model was trained only on those patients that had a final outcome and associated date within the specified date range. As such, we fit the same model but with the inclusion of those patients that were said to be still on the unit at the time of the extract (n = 495), and another that included those patients with clear final outcomes, but no data provided (n = 8).

**Secondary Analyses**

The decision to explore the impact of baseline size was informed by previous studies which have reported an association with mortality in the context of ICU patients with COVID-19. Moreover, we noted that the association of occupancy with mortality risk (originally much larger) decreased in size when we shifted from using an earlier CHESS extract with nearly 1000 fewer patients. We hypothesized that this was due to the backfilling by larger units with lower mortality risk but high occupancy rates, as such we introduced the interaction term to determine the validity of this hypothesis. The resulting model is shown in eTable 3 and the interaction illustrated via eFigure 8. There was no notable interaction between patient age and occupancy (data not shown).

We also explored an alternative way of expressing occupancy, as a proportion of baseline capacity rather than of surge capacity (on the day of admission). Data was drawn from the same sources, and the aforementioned modelling method was applied. Both a linear continuous term and a four factor-level threshold term (based on quartiles, which resulted in thresholds at 70%, 100%, and 150% occupancy) were assessed and are in eTable 5.

**Using effective/additional Age to communicate the mortality risk associated with occupancy**

Spiegelhalter draws on the work of Brenner et al to demonstrate that the presence of a risk factor – in this case ICU operational pressure / occupancy – can be expressed in terms of the number of years lost/gained, providing the following two conditions are filled.[16,17] The first is the standard assumption of proportional hazards in Cox regression; this is implicitly fulfilled by the proportionality of ORs arising from a logistic regression’s parameters. The second condition is that hazard increases exponentially with age (i.e., that each year increases the risk by a fixed percentage), which is the case for all-cause mortality in the population. We adopt this approach and present our primary results via an easily interpretable representation of the additional years of mortality risk associated with the presence of high and low occupancy across patients of different ages. Specifically, we produced three predicted trajectories of mortality risk across an age range of 18 to 99 years using our primary model and solved for intersections of these mortality risk curves to produce effective ages corresponding to equality in risk across the three curves. This is done by first choosing an age and reference curve and then solving for the ages on the other curves corresponding to the other two categories that provide an equal expected risk prediction; these effective ages highlight additional “years of risk” when comparing the occupancy categories. This is clearly illustrated in eFigure 7 and gives rise to Figure 2 in the main text.

**eFigure 1: Application of inclusion and exclusion criteria to raw data.**

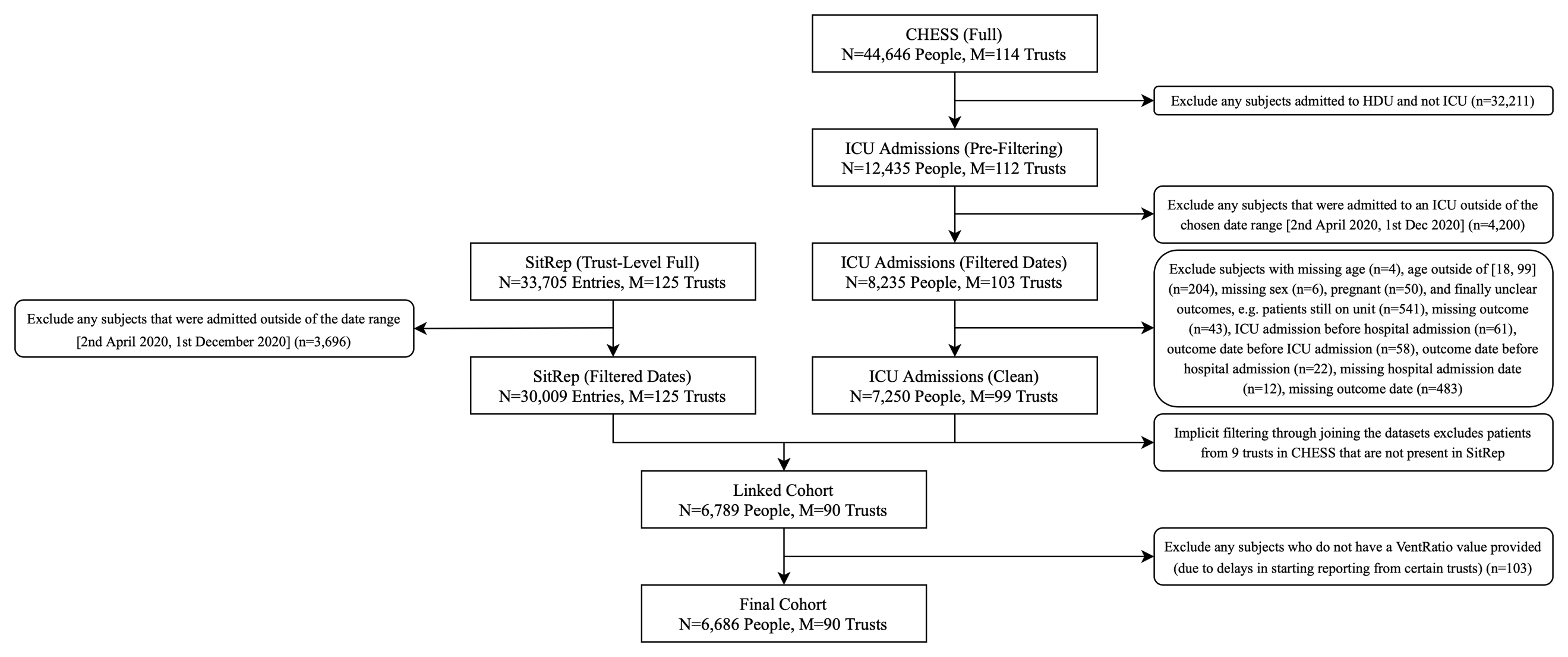

*eFigure 1: Filtering CHESS and SitRep into appropriate cohorts before joining them on date and trust identifier code. More details on how “SitRep (Trust-Level Full)” is reached from raw SitRep data is discussed elsewhere.[1]*

**eFigure 2: Unadjusted mortality rate in the ICU, alongside the total, confirmed COVID and non-COVID national-level occupancies (Top) and percentages of the study cohort in each of the three main ICU occupancy bins used in the primary model (Bottom) across the duration of the study.**

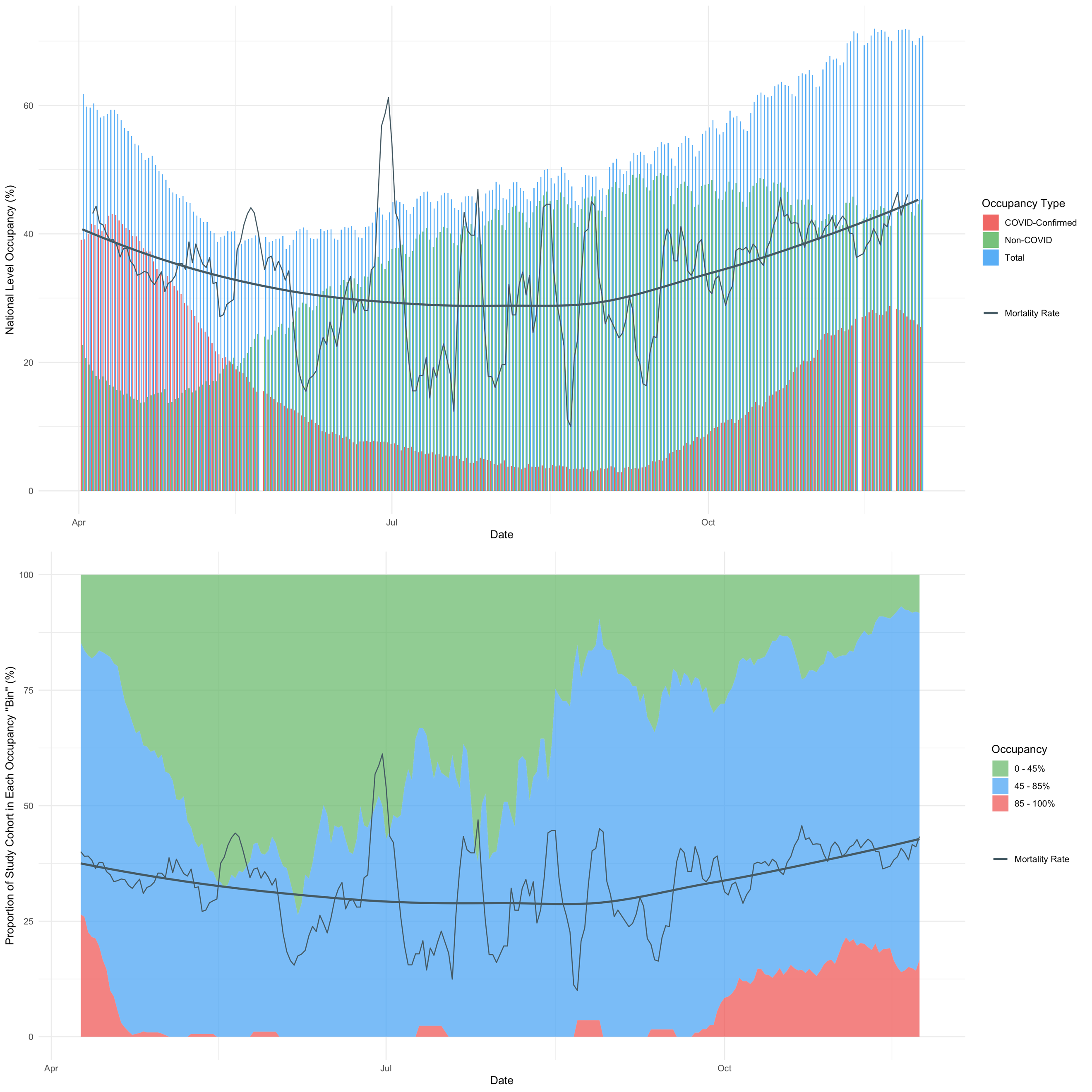

*eFigure 2: Occupancy here is in reference to the number of beds compatible with mechanical ventilation. A loess smoothed mortality curve is shown in black bold alongside the 7-day rolling average of the true daily mortality in the study cohort shown by a thinner black line – Note: CHESS entries were sparse over the summer months and thus there is increased volatility of the mortality statistics during this period.. The COVID and non-COVID occupancies can be seen to cross over in mid-May as hospitals began to admit more non-COVID patients that were initially refused or sent home during the first wave. Previous studies have clearly demonstrated that there is heterogeneity at the local level, and that at 60% national-level occupancy there were several hospitals that were fully saturated. [18]*

**eFigure 3: Trust-level missingness at varying inclusion thresholds.**

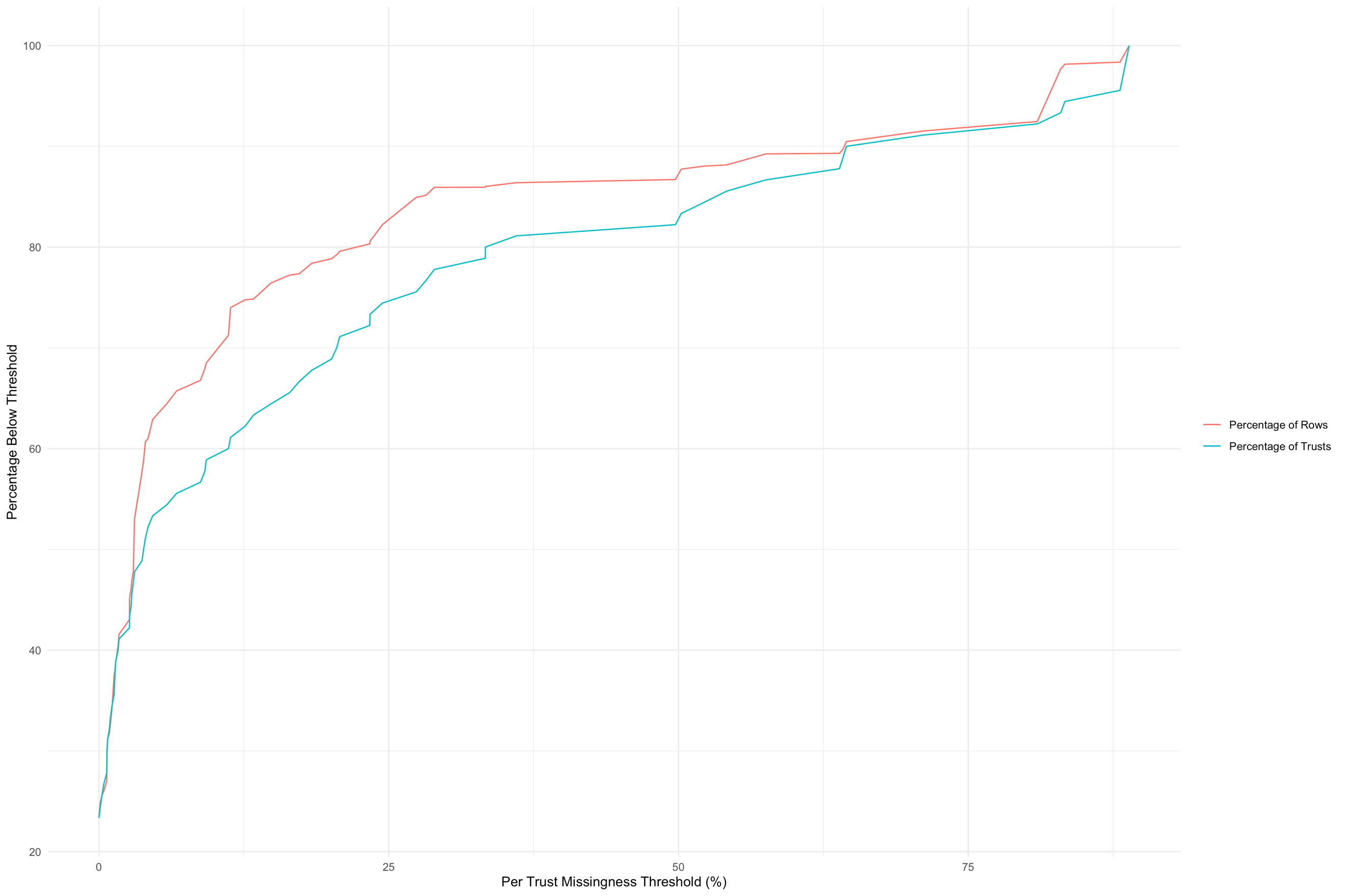

*eFigure 3: Illustrates the percentage of rows and trusts that remain when filtering the full study cohort for varying levels of missingness within each trust. Note that this missingness is calculated as a “pre-cleaning” value; there is no missing data within the modelling cohort. Missing comorbidity indicators are inferred as the absence of that comorbidity; other sources of missingness are excluded before analysis is carried out as illustrated by eFigure 1.*

**eFigure 4: Seven-day rolling averages for Age (Top) and Number of Comorbidities (Bottom) across the duration of the study.**

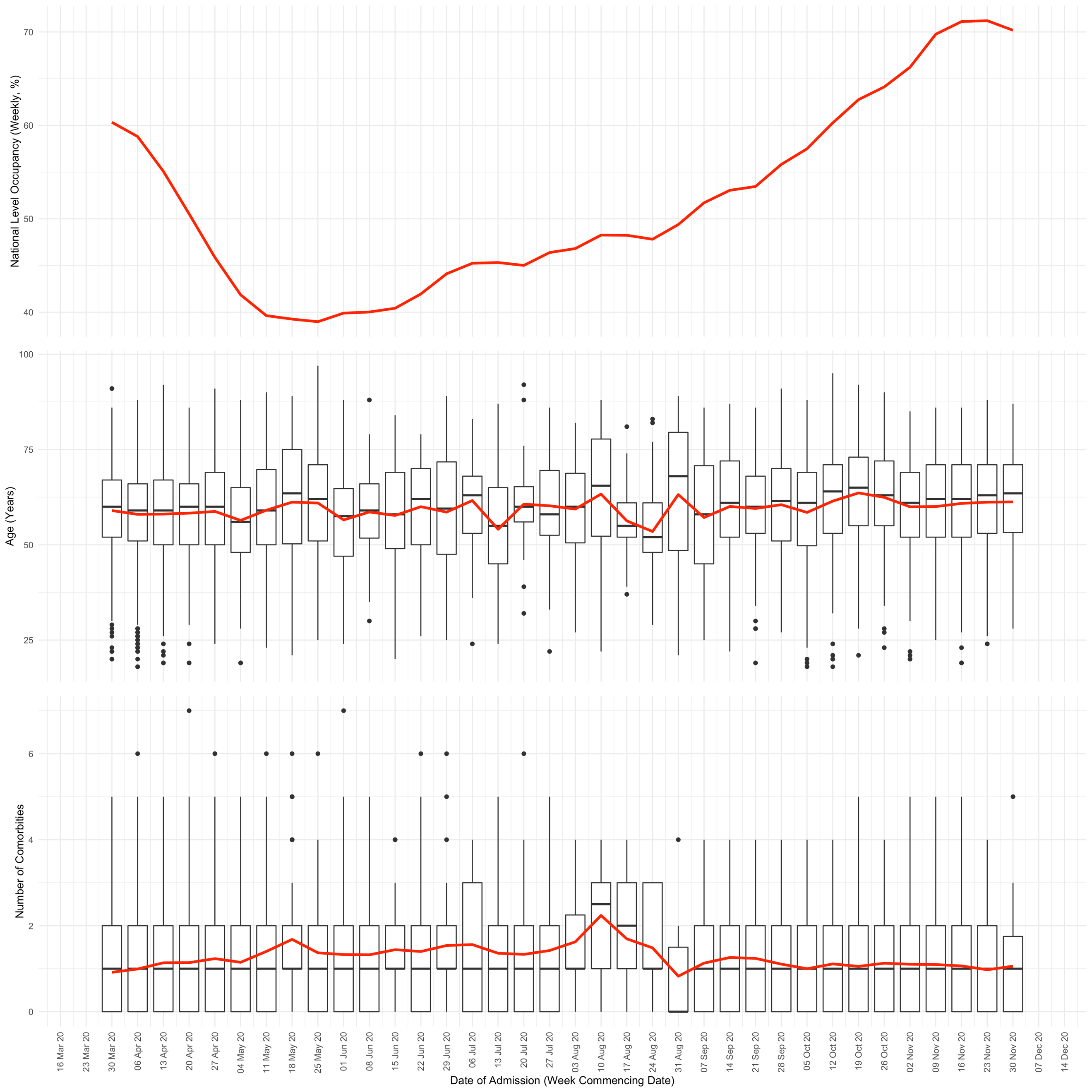

*eFigure 4: By visual inspection the results don’t appear to show any substantial variation, except for a possible increase in comorbidity burden near the end of the first wave and into summer. This results are counter to the hypothesis that younger and more well people were admitted (on average) later in the pandemic (i.e. during periods of lower occupancy), which had it occurred might explain the lower mortality rate. The red line in each part of this figure shows the weekly average value of the relevant y-axis specified quantity. Whilst the box plots in the bottom two sections show the median, IQR and tails of the distribution of patient values within each week.*

**eFigure 5: Marginal posterior densities for linear population level coefficients in the model, for mechanically ventilated bed occupancy binned as described in the main text, comorbidities, sex and week of admission.**

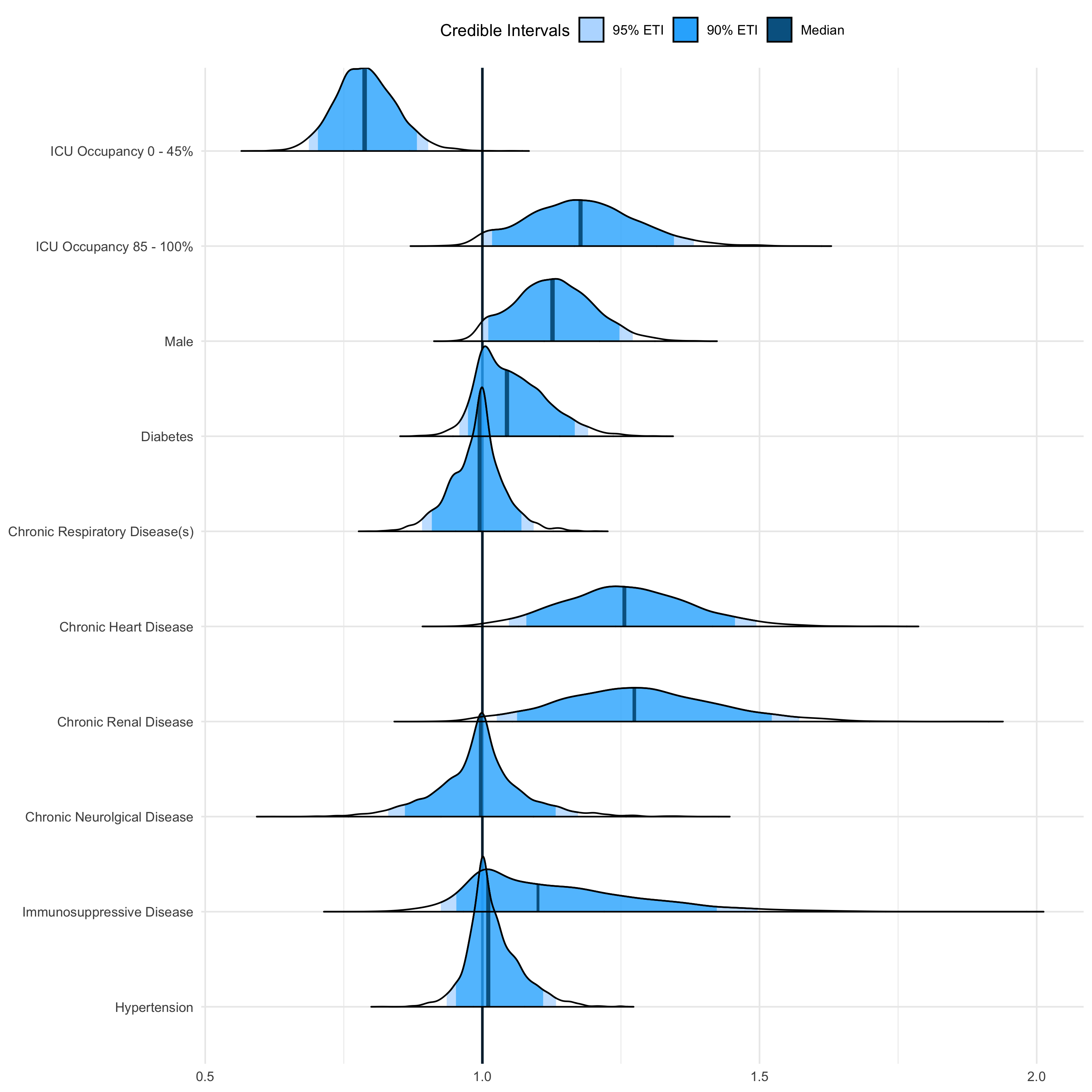

|  | **Posterior Credible Intervals** | | | | |  | **Median** |
| --- | --- | --- | --- | --- | --- | --- | --- |
|  | **95%** | |  | **90%** | |  |  |
|  | Lower | Upper |  | Lower | Upper |  |  |
| **Characteristic** |  |  |  |  |  |  |  |
| Occupancy 0 - 45% | 0.69 | 0.90 |  | 0.70 | 0.88 |  | 0.79 |
| Occupancy 85 - 100% | 1.00 | 1.38 |  | 1.02 | 1.35 |  | 1.18 |
| Male | 1.00 | 1.27 |  | 1.01 | 1.25 |  | 1.13 |
| Diabetes | 0.96 | 1.19 |  | 0.97 | 1.17 |  | 1.04 |
| Chronic Respiratory Disease(s) | 0.89 | 1.09 |  | 0.91 | 1.07 |  | 0.99 |
| Chronic Heart Disease | 1.05 | 1.50 |  | 1.08 | 1.46 |  | 1.26 |
| Chronic Renal Disease | 1.03 | 1.57 |  | 1.06 | 1.52 |  | 1.27 |
| Chronic Neurological Disease | 0.83 | 1.17 |  | 0.86 | 1.13 |  | 1.00 |
| Immunosuppressive Disease | 0.92 | 1.50 |  | 0.95 | 1.42 |  | 1.10 |
| Hypertension | 0.94 | 1.13 |  | 0.95 | 1.11 |  | 1.01 |

*eFigure 5: Female was the reference category for sex, whilst the lack of a comorbidity was the reference category for all of the relevant characteristics.*

**eFigure 6: Marginal posterior densities for group level ethnicity intercepts.**

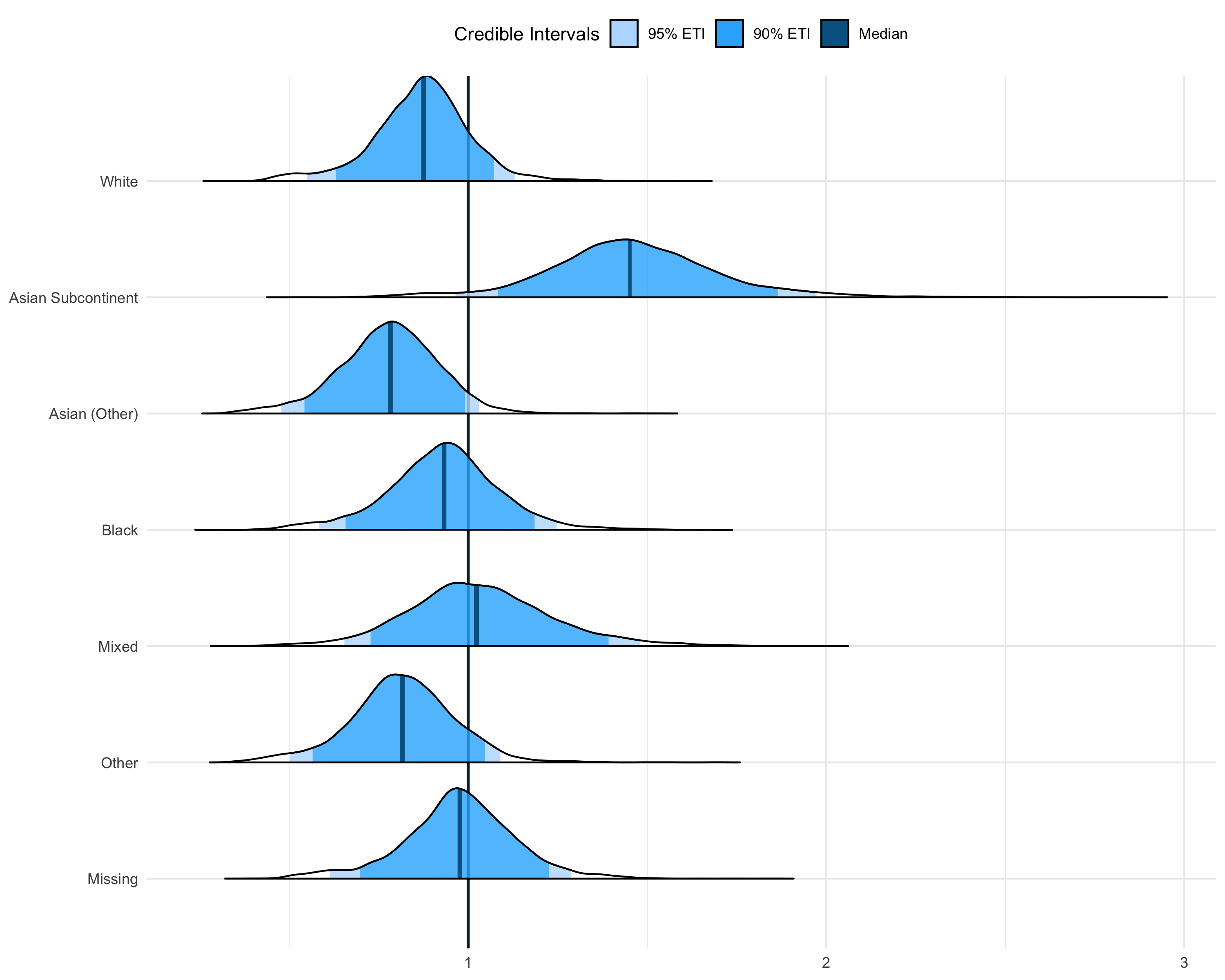

|  | **Posterior Credible Intervals** | | | | |  | **Median** |
| --- | --- | --- | --- | --- | --- | --- | --- |
|  | **95%** | |  | **90%** | |  |  |
|  | Lower | Upper |  | Lower | Upper |  |  |
| **Characteristic** |  |  |  |  |  |  |  |
| White | 0.55 | 1.13 |  | 0.63 | 1.07 |  | 0.88 |
| Asian Subcontinent | 0.96 | 1.98 |  | 1.08 | 1.87 |  | 1.45 |
| Asian (Other) | 0.48 | 1.03 |  | 0.54 | 0.99 |  | 0.78 |
| Black | 0.58 | 1.25 |  | 0.66 | 1.19 |  | 0.93 |
| Mixed | 0.65 | 1.48 |  | 0.72 | 1.39 |  | 1.02 |
| Other | 0.50 | 1.09 |  | 0.56 | 1.05 |  | 0.82 |
| Missing | 0.61 | 1.29 |  | 0.70 | 1.23 |  | 0.98 |

*eFigure 6: It can be seen that Asian Subcontinent (including Indian, Bangladeshi and Pakistani ethnicities) is the only significant group intercept according to the 90% credible intervals.*

**eFigure 7: Predicted mortality risk for each of the three categorical levels of occupancy, illustrating how effective ages are calculated for Figure 2.**

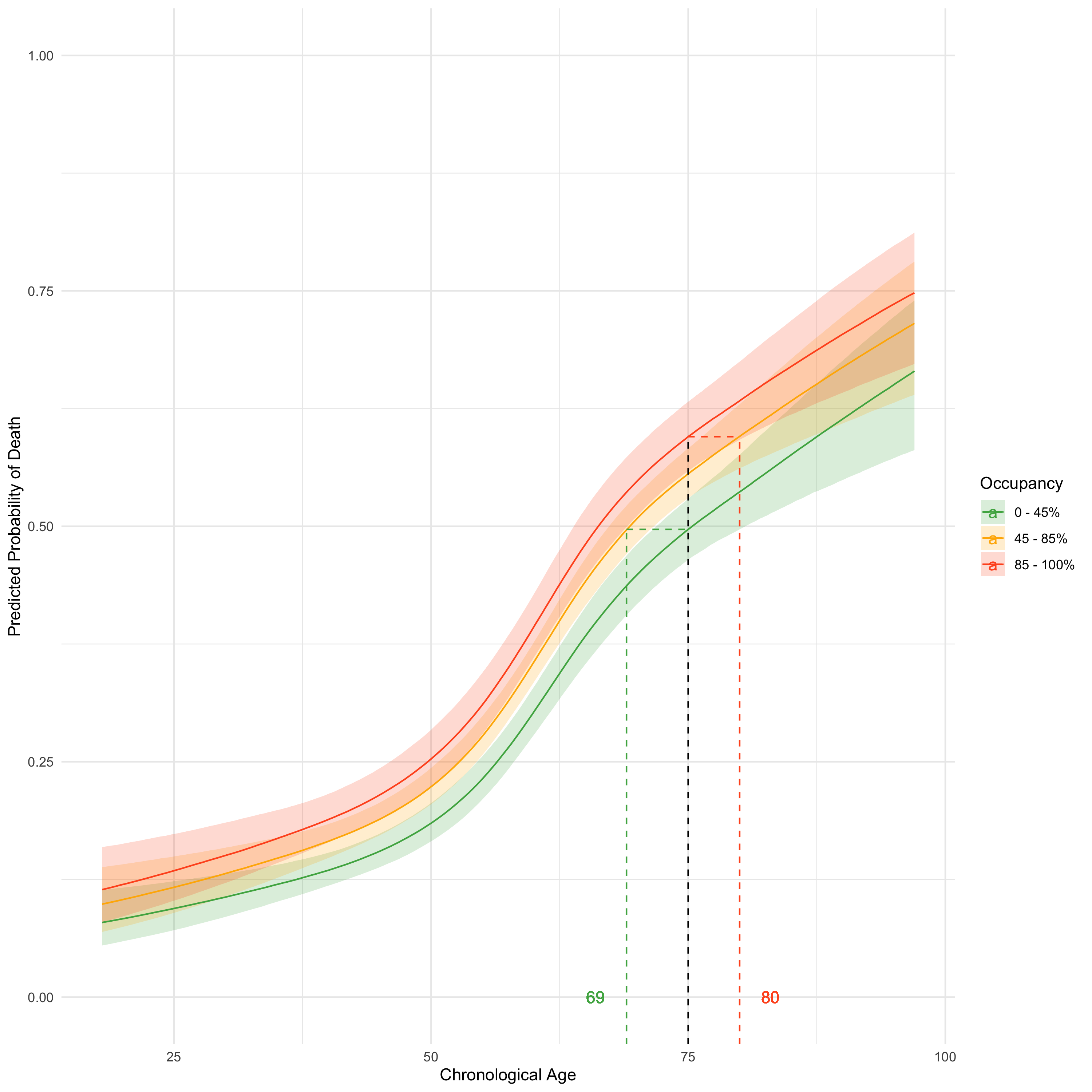

*eFigure 7: The predicted mortality curves arising from predictions made by the primary model across a granular range of age values for a white male patient is shown alongside 95% credible intervals in a ribbon either side of the median line. The black dotted line intersects all three curves; the 0 – 45% and 85 – 100% occupancy curve y-value probabilities can then be used to solve back onto the reference curve to determine effective ages of equal risk to the chosen age under reference 45 – 85% occupancy, shown by the corresponding green and red dotted lines respectively.*

**eFigure 8: The interaction between the baseline availability of beds supporting mechanical ventilation, and occupancy on the day of ICU admission.**

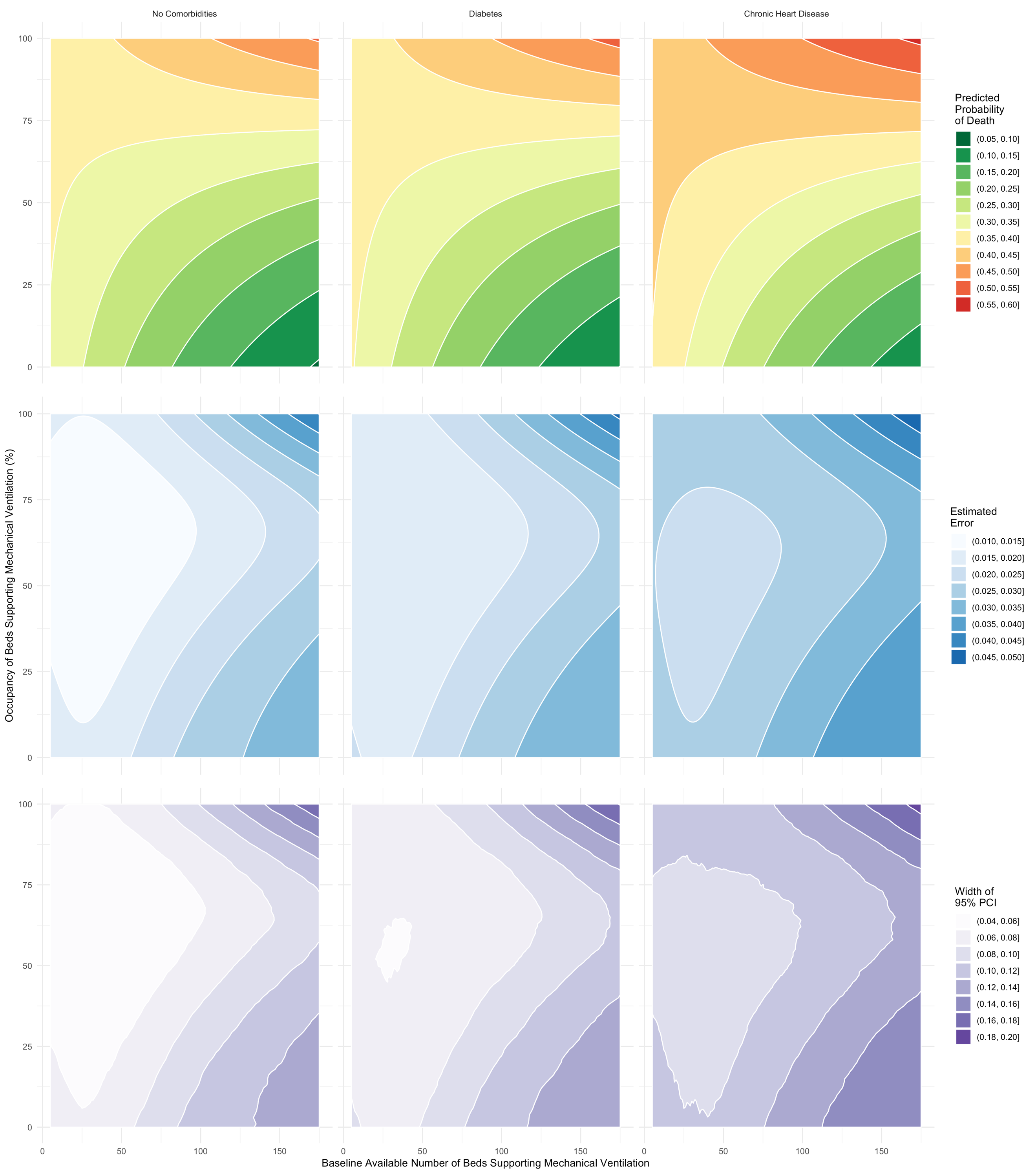

*eFigure 8: Three illustrative contour plots for risk of mortality based on the model described in eTable 3 (using a white male 60-year-old patient in each), varying only pre-pandemic size and occupancy (with an interaction term): (left) patient with no co-morbidities; (middle) patient with diabetes; (right) patient with chronic heart disease. In all three plots the variability in mortality risk associated with different degrees of bed occupancy on the day of admission increases with pre-pandemic size of the unit. The estimated errors associated with the expected predictions across all sampled model parameters are shown in the second row of plots, and the 95% credible intervals (empirical across all samples) are illustrated in the third row.*

**eTable 1: Marginal posterior densities for occupancy under sensitivity analyses.**

|  | **Posterior Credible Intervals** | | | | |  | **Median** |
| --- | --- | --- | --- | --- | --- | --- | --- |
|  | **95%** | |  | **90%** | |  |  |
|  | Lower | Upper |  | Lower | Upper |  |  |
| **Primary** | | | | | | | |
| 0 - 45% | 0.69 | 0.90 |  | 0.70 | 0.88 |  | 0.79 |
| 85 - 100% | 1.00 | 1.38 |  | 1.02 | 1.35 |  | 1.18 |
| **Minimal / Unadjusted** | | | | | | | |
| 0 - 45% | 0.70 | 0.90 |  | 0.71 | 0.88 |  | 0.79 |
| 85 - 100% | 1.00 | 1.33 |  | 1.01 | 1.30 |  | 1.14 |
| **With Week of Admission (Continuous)** | | | | | | | |
| 0 - 45% | 0.68 | 0.89 |  | 0.69 | 0.87 |  | 0.77 |
| 85 - 100% | 0.99 | 1.33 |  | 1.00 | 1.30 |  | 1.14 |
| **With Time to ICU in Days (Continuous)** | | | | | | | |
| 0 - 45% | 0.69 | 0.91 |  | 0.70 | 0.89 |  | 0.79 |
| 85 - 100% | 1.00 | 1.37 |  | 1.02 | 1.34 |  | 1.17 |
| **With Chronic Liver Disease (Indicator)** | | | | | | | |
| 0 - 45% | 0.69 | 0.91 |  | 0.70 | 0.89 |  | 0.79 |
| 85 - 100% | 1.00 | 1.37 |  | 1.02 | 1.34 |  | 1.18 |
| **75% Missingness Threshold** | | | | | | | |
| 0 - 45% | 0.73 | 0.99 |  | 0.75 | 0.97 |  | 0.85 |
| 85 - 100% | 1.02 | 1.47 |  | 1.05 | 1.43 |  | 1.23 |
| **50% Missingness Threshold** | | | | | | | |
| 0 - 45% | 0.72 | 0.98 |  | 0.74 | 0.96 |  | 0.84 |
| 85 - 100% | 1.03 | 1.49 |  | 1.07 | 1.46 |  | 1.26 |
| **25% Missingness Threshold** | | | | | | | |
| 0 - 45% | 0.70 | 0.96 |  | 0.72 | 0.94 |  | 0.82 |
| 85 - 100% | 1.02 | 1.46 |  | 1.05 | 1.43 |  | 1.23 |
| **With Obesity (Group-Level Effect)** | | | | | | | |
| 0 - 45% | 0.68 | 0.90 |  | 0.70 | 0.88 |  | 0.78 |
| 85 - 100% | 0.99 | 1.36 |  | 1.00 | 1.33 |  | 1.15 |
| **With Baseline Bed Availability (Continuous)** | | | | | | | |
| 0 - 45% | 0.68 | 0.89 |  | 0.70 | 0.88 |  | 0.78 |
| 85 - 100% | 1.00 | 1.36 |  | 1.01 | 1.33 |  | 1.16 |
| **With Region (Group-Level Effect)** | | | | | | | |
| 0 - 45% | 0.71 | 0.95 |  | 0.73 | 0.93 |  | 0.82 |
| 85 - 100% | 1.01 | 1.40 |  | 1.04 | 1.36 |  | 1.19 |
| **With Trust (Group-Level Effect)** | | | | | | | |
| 0 - 45% | 0.66 | 0.92 |  | 0.68 | 0.89 |  | 0.77 |
| 85 - 100% | 1.02 | 1.46 |  | 1.05 | 1.42 |  | 1.22 |
| **With Dexamethasone Treatment (Indicator)** | | | | | | | |
| 0 - 45% | 0.68 | 0.88 |  | 0.69 | 0.87 |  | 0.77 |
| 85 - 100% | 1.00 | 1.35 |  | 1.01 | 1.32 |  | 1.15 |
| **Data Including Patients that were Still on the Unit as of 22nd December (Add. n = 495)** | | | | | | | |
| 0 - 45% | 0.75 | 0.98 |  | 0.76 | 0.96 |  | 0.86 |
| 85 - 100% | 1.08 | 1.46 |  | 1.11 | 1.43 |  | 1.26 |
| **Data Including Patients that were Missing a Final Outcome Date * (Add. n = 8)** | | | | | | | |
| 0 - 45% | 0.69 | 0.90 |  | 0.70 | 0.88 |  | 0.79 |
| 85 - 100% | 1.00 | 1.38 |  | 1.01 | 1.35 |  | 1.17 |

*Legend: Note that missingness thresholds define inclusion of each trust in the dataset, as shown in eFigure 2; i.e. only trusts with maximum 25% missingness were included in the data used to fit the model for the “25% Missingness Threshold” sensitivity. 2,497 patients were admitted after June 16^th^, and so were presumed treated with Dexamethasone, 4,244 patients were admitted before this date and thus were not. * Patients that were still on ICU were not added, only those with clearly resolved outcomes but no date.*

**eTable 2: Marginal posterior densities for mortality ORs when occupancy is utilised at each individual’s final outcome date, rather than their date of admission to ICU.**

|  | **Posterior Credible Intervals** | | | | |  | **Median** |
| --- | --- | --- | --- | --- | --- | --- | --- |
|  | **95%** | |  | **90%** | |  |  |
|  | Lower | Upper |  | Lower | Upper |  |  |
| **Occupancy on Outcome Date, Categorical** | | | | | | | |
| Occupancy 0 - 45% | 0.58 | 0.74 |  | 0.59 | 0.73 |  | 0.65 |
| Occupancy 85 - 100% | 0.99 | 1.43 |  | 1.00 | 1.38 |  | 1.17 |
| Male | 0.99 | 1.26 |  | 1.00 | 1.24 |  | 1.11 |
| Diabetes | 0.97 | 1.23 |  | 0.99 | 1.21 |  | 1.08 |
| Chronic Respiratory Disease(s) | 0.87 | 1.09 |  | 0.90 | 1.06 |  | 0.99 |
| Chronic Heart Disease | 1.03 | 1.47 |  | 1.06 | 1.43 |  | 1.24 |
| Chronic Renal Disease | 1.00 | 1.53 |  | 1.03 | 1.48 |  | 1.24 |
| Chronic Neurological Disease | 0.80 | 1.18 |  | 0.84 | 1.13 |  | 0.99 |
| Immunosuppressive Disease | 0.93 | 1.52 |  | 0.95 | 1.44 |  | 1.11 |
| Hypertension | 0.96 | 1.18 |  | 0.97 | 1.16 |  | 1.04 |
| **Occupancy on Outcome Date, Linear Proportional** | | | | | | | |
| Continuous Linear Variable | 2.33 | 3.83 |  | 2.42 | 3.68 |  | 2.98 |
| Male | 1.00 | 1.27 |  | 1.02 | 1.25 |  | 1.13 |
| Diabetes | 0.97 | 1.23 |  | 0.98 | 1.20 |  | 1.08 |
| Chronic Respiratory Disease(s) | 0.88 | 1.08 |  | 0.90 | 1.07 |  | 0.99 |
| Chronic Heart Disease | 1.01 | 1.46 |  | 1.05 | 1.43 |  | 1.23 |
| Chronic Renal Disease | 1.00 | 1.54 |  | 1.03 | 1.48 |  | 1.24 |
| Chronic Neurological Disease | 0.81 | 1.18 |  | 0.83 | 1.13 |  | 0.99 |
| Immunosuppressive Disease | 0.92 | 1.51 |  | 0.95 | 1.45 |  | 1.11 |
| Hypertension | 0.95 | 1.18 |  | 0.97 | 1.15 |  | 1.04 |

**eTable 3: Marginal posterior densities for mortality ORs including baseline bed availability and interaction with occupancy.**

|  | **Posterior Credible Intervals** | | | | |  | **Median** |
| --- | --- | --- | --- | --- | --- | --- | --- |
|  | **95%** | |  | **90%** | |  |  |
|  | Lower | Upper |  | Lower | Upper |  |  |
| **With Baseline Bed Availability and Linear Occupancy** | | | | | | | |
| Baseline Bed Availability (Per 25 beds) | 0.95 | 1.01 |  | 0.95 | 1.01 |  | 0.98 |
| Occupancy (Linear) | 1.29 | 2.21 |  | 1.36 | 2.13 |  | 1.72 |
| Male | 1.00 | 1.27 |  | 1.02 | 1.25 |  | 1.13 |
| Diabetes | 0.96 | 1.18 |  | 0.97 | 1.16 |  | 1.04 |
| Chronic Respiratory Disease(s) | 0.89 | 1.09 |  | 0.91 | 1.07 |  | 1.00 |
| Chronic Heart Disease | 1.05 | 1.50 |  | 1.08 | 1.46 |  | 1.27 |
| Chronic Renal Disease | 1.01 | 1.57 |  | 1.04 | 1.51 |  | 1.27 |
| Chronic Neurological Disease | 0.83 | 1.18 |  | 0.86 | 1.13 |  | 1.00 |
| Immunosuppressive Disease | 0.92 | 1.50 |  | 0.95 | 1.43 |  | 1.09 |
| Hypertension | 0.93 | 1.12 |  | 0.95 | 1.11 |  | 1.01 |
| **With Baseline Bed Availability, Linear Occupancy and an Interaction Term** | | | | | | | |
| Baseline Bed Availability (Per 25 beds) | 0.70 | 0.88 |  | 0.72 | 0.86 |  | 0.78 |
| Occupancy (Linear) | 0.87 | 1.39 |  | 0.91 | 1.30 |  | 1.01 |
| Interaction Term | 1.01 | 1.02 |  | 1.01 | 1.02 |  | 1.01 |
| Male | 0.99 | 1.26 |  | 1.00 | 1.23 |  | 1.11 |
| Diabetes | 0.96 | 1.18 |  | 0.98 | 1.15 |  | 1.03 |
| Chronic Respiratory Disease(s) | 0.90 | 1.08 |  | 0.92 | 1.06 |  | 1.00 |
| Chronic Heart Disease | 1.02 | 1.50 |  | 1.06 | 1.46 |  | 1.25 |
| Chronic Renal Disease | 1.00 | 1.56 |  | 1.03 | 1.50 |  | 1.25 |
| Chronic Neurological Disease | 0.83 | 1.13 |  | 0.87 | 1.10 |  | 1.00 |
| Immunosuppressive Disease | 0.93 | 1.45 |  | 0.96 | 1.37 |  | 1.05 |
| Hypertension | 0.94 | 1.11 |  | 0.95 | 1.09 |  | 1.01 |

**eTable 4: Comparison of prior choices via their effect on primary model OR estimates.**

| **Prior Type** |  | **Posterior Credible Intervals** | | | | |  | **Median** |
| --- | --- | --- | --- | --- | --- | --- | --- | --- |
|  |  | **95%** | |  | **90%** | |  |  |
|  |  | Lower | Upper |  | Lower | Upper |  |  |
| **Hierarchical Shrinkage, DF = 1** |  |  |  |  |  |  |  |  |
| Occupancy 0 - 45% |  | 0.69 | 0.90 |  | 0.70 | 0.88 |  | 0.79 |
| Occupancy 85 - 100% |  | 1.00 | 1.38 |  | 1.02 | 1.34 |  | 1.17 |
| Male |  | 1.00 | 1.27 |  | 1.01 | 1.25 |  | 1.13 |
| Diabetes |  | 0.95 | 1.19 |  | 0.97 | 1.16 |  | 1.04 |
| Chronic Respiratory Disease(s) |  | 0.89 | 1.10 |  | 0.90 | 1.07 |  | 0.99 |
| Chronic Heart Disease |  | 1.05 | 1.50 |  | 1.09 | 1.46 |  | 1.26 |
| Chronic Renal Disease |  | 1.03 | 1.56 |  | 1.07 | 1.52 |  | 1.28 |
| Chronic Neurological Disease |  | 0.83 | 1.18 |  | 0.86 | 1.14 |  | 1.00 |
| Immunosuppressive Disease |  | 0.92 | 1.51 |  | 0.95 | 1.43 |  | 1.10 |
| Hypertension |  | 0.93 | 1.13 |  | 0.95 | 1.11 |  | 1.01 |
| **Hierarchical Shrinkage, DF = 3** |  |  |  |  |  |  |  |  |
| Occupancy 0 - 45% |  | 0.69 | 0.90 |  | 0.70 | 0.89 |  | 0.79 |
| Occupancy 85 - 100% |  | 1.00 | 1.37 |  | 1.02 | 1.34 |  | 1.18 |
| Male |  | 1.00 | 1.28 |  | 1.01 | 1.25 |  | 1.13 |
| Diabetes |  | 0.96 | 1.19 |  | 0.97 | 1.17 |  | 1.05 |
| Chronic Respiratory Disease(s) |  | 0.88 | 1.10 |  | 0.90 | 1.07 |  | 0.99 |
| Chronic Heart Disease |  | 1.06 | 1.49 |  | 1.09 | 1.45 |  | 1.27 |
| Chronic Renal Disease |  | 1.02 | 1.55 |  | 1.06 | 1.50 |  | 1.27 |
| Chronic Neurological Disease |  | 0.82 | 1.18 |  | 0.85 | 1.14 |  | 1.00 |
| Immunosuppressive Disease |  | 0.92 | 1.50 |  | 0.95 | 1.42 |  | 1.10 |
| Hypertension |  | 0.93 | 1.13 |  | 0.95 | 1.11 |  | 1.01 |
| **Hierarchical Shrinkage, DF = 5** |  |  |  |  |  |  |  |  |
| Occupancy 0 - 45% |  | 0.69 | 0.90 |  | 0.70 | 0.88 |  | 0.79 |
| Occupancy 85 - 100% |  | 1.01 | 1.38 |  | 1.03 | 1.34 |  | 1.18 |
| Male |  | 1.00 | 1.27 |  | 1.01 | 1.25 |  | 1.13 |
| Diabetes |  | 0.96 | 1.19 |  | 0.97 | 1.16 |  | 1.05 |
| Chronic Respiratory Disease(s) |  | 0.89 | 1.09 |  | 0.91 | 1.07 |  | 0.99 |
| Chronic Heart Disease |  | 1.05 | 1.49 |  | 1.08 | 1.45 |  | 1.26 |
| Chronic Renal Disease |  | 1.02 | 1.55 |  | 1.06 | 1.50 |  | 1.27 |
| Chronic Neurological Disease |  | 0.81 | 1.18 |  | 0.84 | 1.14 |  | 1.00 |
| Immunosuppressive Disease |  | 0.92 | 1.51 |  | 0.95 | 1.43 |  | 1.12 |
| Hypertension |  | 0.94 | 1.13 |  | 0.95 | 1.11 |  | 1.01 |
| **Hierarchical Shrinkage, DF = 7** |  |  |  |  |  |  |  |  |
| Occupancy 0 - 45% |  | 0.69 | 0.90 |  | 0.70 | 0.88 |  | 0.79 |
| Occupancy 85 - 100% |  | 1.01 | 1.38 |  | 1.04 | 1.34 |  | 1.18 |
| Male |  | 1.00 | 1.27 |  | 1.02 | 1.25 |  | 1.13 |
| Diabetes |  | 0.96 | 1.20 |  | 0.97 | 1.17 |  | 1.05 |
| Chronic Respiratory Disease(s) |  | 0.89 | 1.09 |  | 0.91 | 1.07 |  | 0.99 |
| Chronic Heart Disease |  | 1.06 | 1.50 |  | 1.09 | 1.46 |  | 1.26 |
| Chronic Renal Disease |  | 1.04 | 1.54 |  | 1.08 | 1.50 |  | 1.27 |
| Chronic Neurological Disease |  | 0.81 | 1.18 |  | 0.85 | 1.14 |  | 1.00 |
| Immunosuppressive Disease |  | 0.92 | 1.52 |  | 0.95 | 1.44 |  | 1.11 |
| Hypertension |  | 0.93 | 1.13 |  | 0.94 | 1.11 |  | 1.01 |
| **Standard Normal** |  |  |  |  |  |  |  |  |
| Occupancy 0 - 45% |  | 0.69 | 0.89 |  | 0.70 | 0.87 |  | 0.78 |
| Occupancy 85 - 100% |  | 1.05 | 1.42 |  | 1.08 | 1.39 |  | 1.22 |
| Male |  | 1.03 | 1.30 |  | 1.05 | 1.28 |  | 1.16 |
| Diabetes |  | 0.95 | 1.22 |  | 0.97 | 1.20 |  | 1.08 |
| Chronic Respiratory Disease(s) |  | 0.87 | 1.12 |  | 0.89 | 1.10 |  | 0.99 |
| Chronic Heart Disease |  | 1.09 | 1.52 |  | 1.13 | 1.49 |  | 1.29 |
| Chronic Renal Disease |  | 1.09 | 1.61 |  | 1.12 | 1.56 |  | 1.32 |
| Chronic Neurological Disease |  | 0.77 | 1.23 |  | 0.80 | 1.19 |  | 0.97 |
| Immunosuppressive Disease |  | 0.93 | 1.69 |  | 0.97 | 1.60 |  | 1.25 |
| Hypertension |  | 0.92 | 1.15 |  | 0.93 | 1.13 |  | 1.02 |

**eTable 5: Marginal posterior densities for mortality ORs when occupancy is defined as a ratio relative to baseline capacity.**

|  | **Posterior Credible Intervals** | | | | |  | **Median** |
| --- | --- | --- | --- | --- | --- | --- | --- |
|  | **95%** | |  | **90%** | |  |  |
|  | Lower | Upper |  | Lower | Upper |  |  |
| **With Occupancy Relative to Baseline Binned in Groups** | | | | | | | |
| Occupancy Rel. to Baseline (70 - 100%) | 0.88 | 1.09 |  | 0.90 | 1.08 |  | 1.00 |
| Occupancy Rel. to Baseline (100 - 150%) | 1.12 | 1.47 |  | 1.15 | 1.44 |  | 1.29 |
| Occupancy Rel. to Baseline (>150%) | 0.97 | 1.38 |  | 0.99 | 1.33 |  | 1.12 |
| Male | 1.00 | 1.27 |  | 1.01 | 1.25 |  | 1.13 |
| Diabetes | 0.96 | 1.20 |  | 0.97 | 1.17 |  | 1.05 |
| Chronic Respiratory Disease(s) | 0.89 | 1.10 |  | 0.91 | 1.08 |  | 1.00 |
| Chronic Heart Disease | 1.06 | 1.52 |  | 1.09 | 1.48 |  | 1.28 |
| Chronic Renal Disease | 1.02 | 1.56 |  | 1.05 | 1.52 |  | 1.28 |
| Chronic Neurological Disease | 0.82 | 1.16 |  | 0.85 | 1.12 |  | 1.00 |
| Immunosuppressive Disease | 0.92 | 1.48 |  | 0.95 | 1.40 |  | 1.07 |
| Hypertension | 0.93 | 1.13 |  | 0.95 | 1.11 |  | 1.01 |
| **With Linear Occupancy Relative to Baseline** | | | | | | | |
| Occupancy Relative to Baseline (Linear) | 1.13 | 1.43 |  | 1.15 | 1.41 |  | 1.27 |
| Male | 1.00 | 1.27 |  | 1.02 | 1.25 |  | 1.13 |
| Diabetes | 0.96 | 1.19 |  | 0.97 | 1.16 |  | 1.04 |
| Chronic Respiratory Disease(s) | 0.89 | 1.09 |  | 0.91 | 1.07 |  | 1.00 |
| Chronic Heart Disease | 1.06 | 1.52 |  | 1.10 | 1.49 |  | 1.28 |
| Chronic Renal Disease | 1.04 | 1.58 |  | 1.08 | 1.52 |  | 1.29 |
| Chronic Neurological Disease | 0.81 | 1.17 |  | 0.84 | 1.12 |  | 0.99 |
| Immunosuppressive Disease | 0.92 | 1.48 |  | 0.95 | 1.41 |  | 1.08 |
| Hypertension | 0.93 | 1.12 |  | 0.94 | 1.10 |  | 1.01 |
